# Survivors of SARS-CoV-2 Infection Show Neuropsychiatric Sequelae Measured by Surveys, Neurocognitive Testing, and Magnetic Resonance Imaging: Preliminary Results

**DOI:** 10.1101/2021.08.23.21256078

**Authors:** Laura M. Hack, Jacob Brawer, Megan Chesnut, Xue Zhang, Max Wintermark, Bin Jiang, Philip Grant, Hector Bonilla, Patrick Stetz, Jerome A. Yesavage, Aruna Subramanian, Leanne M. Williams

## Abstract

A significant number of individuals experience physical, cognitive, and mental health symptoms in the months after acute infection with SARS-CoV-2, the virus that causes COVID-19. This study assessed depressive and anxious symptoms, cognition, and brain structure and function in participants with symptomatic COVID-19 confirmed by PCR testing (n=100) approximately three months following infection, leveraging self-report questionnaires, objective neurocognitive testing, and structural and functional neuroimaging data. Preliminary results demonstrated that over 1/5 of our cohort endorsed clinically significant depressive and/or anxious symptoms, and >40% of participants had cognitive impairment on objective testing across multiple domains, consistent with ‘brain-fog’. While depression and one domain of quality of life (physical functioning) were significantly different between hospitalized and non-hospitalized participants, anxiety, cognitive impairment, and most domains of functioning were not, suggesting that the severity of SARS-CoV-2 infection does not necessarily relate to the severity of neuropsychiatric outcomes and impaired functioning in the months after infection. Furthermore, we found that the majority of participants in a subset of our cohort who completed structural and functional neuroimaging (n=15) had smaller olfactory bulbs and sulci in conjunction with anosmia. We also showed that this subset of participants had dysfunction in attention network functional connectivity and ventromedial prefrontal cortex seed-based functional connectivity. These functional imaging dysfunctions have been observed previously in depression and correlated with levels of inflammation. Our results support and extend previous findings in the literature concerning the neuropsychiatric sequelae associated with long COVID. Ongoing data collection and analyses within this cohort will allow for a more comprehensive understanding of the longitudinal relationships between neuropsychiatric symptoms, neurocognitive performance, brain structure and function, and inflammatory and immune profiles.

## Introduction

Following recovery from acute infection with SARS-CoV-2, the virus that causes COVID-19, between 22% and 44% of patients report psychiatric symptoms, including depression, anxiety, and/or post-traumatic stress disorder (PTSD)^1-3^. For those with pre-existing mental health conditions, COVID-19 has been shown to increase psychiatric symptoms^4^. There is also an elevated risk of a first psychiatric diagnosis in the 14–90-day period post-infection compared to other viral infections^3^. Furthermore, these post-infection impacts have been observed to increase risk of suicide^5^.

It is known that exposure to viruses can acutely lead to depressive and anxious symptoms through the direct effect of peripheral inflammatory cytokines on the brain^6-8^. In animal models, injection of viruses that activate the immune system have been found to induce sickness behavior that resembles depression^7^. SARS-CoV-2 creates an excessive detrimental immune effect in some individuals involving marked elevation in inflammatory cytokines^9^, and there is evidence that this inflammation continues at a lower level in long COVID patients for at least 8 months following infection, along with other immunological abnormalities^10^. A subset of depressed patients have been distinguished by a correlation between low-level inflammation assayed by the acute phase reactant C-reactive protein (CRP)^11^ and disrupted brain circuits assessed by functional MRI^12-14^. To our knowledge, no published studies to date have assessed functional neuroimaging in post-COVID-19 patients.

Cognitive deficits associated with SARS-CoV-2 infection, or ‘brain fog’, ‘cognitive fog’, or ‘COVID brain’, are debilitating and potentially long-lasting neuropsychiatric symptoms that have been found using both self-report questionnaires^15^ and objective testing^16^. While there is no official definition, ‘brain fog’ may involve confusion, short-term memory deficits, difficulty with concentration, slowed processing speed, and mental fatigue. Of note, there is evidence that SARS-CoV-2 is capable of neuroinvasion^17^, and other neurotropic viruses have been associated with long-term cognitive decline and the development of dementia^18^. One potential pathway of neuroinvasion is through the olfactory nerve and bulb. A recent meta-analysis suggests abnormalities in the olfactory bulb are the most common structural neuroimaging findings following COVID-19 infection^19^. Furthermore, structural abnormalities in the olfactory bulb have been associated with anosmia (a partial or complete loss of smell)^20, 21^, and there has been speculation in the literature that olfactory dysfunction in COVID-19 may increase risk of future dementia^22^.

In this report, at approximately three months following COVID-19 diagnosis, we aimed to 1) characterize the prevalence and severity of psychiatric symptoms and neurocognitive deficits; 2) assess structural olfactory abnormalities; and 3) identify neural circuit dysfunction.

## Methods

### Infection Recovery in SARS-CoV-2 (IRIS) Neurostudy cohort

We enrolled 100 participants who were diagnosed with COVID-19 due to symptoms and confirmed by PCR testing from a convenience sample of participants presenting to Stanford^23^. Inclusion criteria for participation in the study were as follows: 1) willing and able to provide written informed consent, or with a legal representative who can provide informed consent; 2) age ≥18 years; and 3) history of symptomatic COVID-19 infection confirmed by PCR test. Exclusion criteria included active drug or alcohol abuse that, in the investigators’ opinion, could prevent compliance with study procedures or confound the analysis of study endpoints. Participants who presented contraindications to MRI scanning were given the opportunity to complete all other study assessments. Institutional approval for the study was obtained through the Stanford Institutional Review Board. Prior to enrollment, written informed consent was obtained. In this longitudinal study, we sought to enroll participants for a first assessment at about three months after COVID-19 diagnosis and reassess them at 6 and 12 months after diagnosis. Due to the challenges of initiating a study during the pandemic, some participants had their first study visit earlier or later than three months. **Fig. 1** shows the time distribution of the first study visit. For the purposes of the analyses presented here, the first time point collected for all participants was combined.

**Fig. 1.**
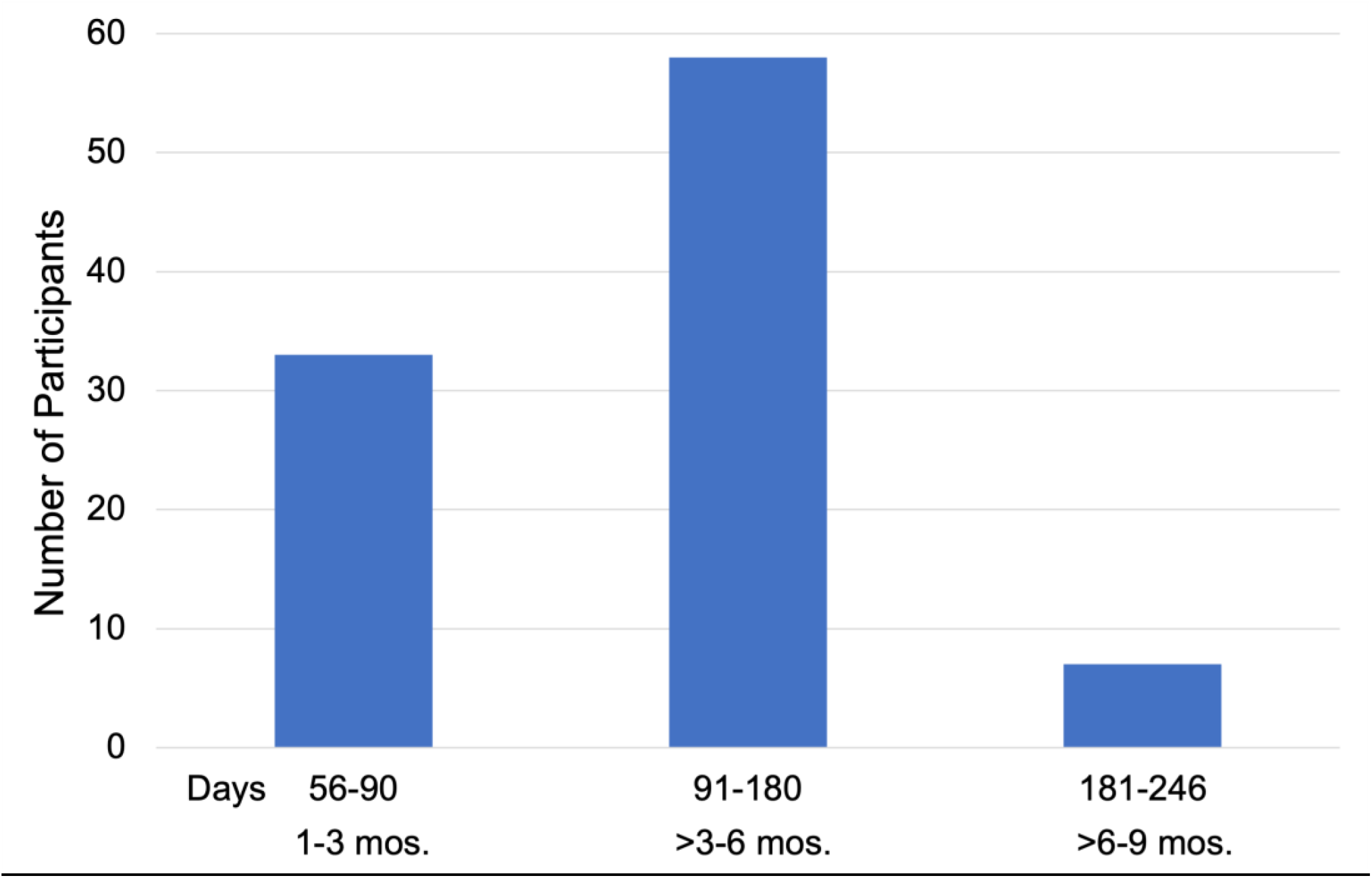
Number of participants who had their first study visit within 1-3 months, >3-6 months, and >6-9 months after COVID-19 diagnosis.

### Demographics, Medical History, and Physical Function

At an initial visit, demographics were collected, including age, biological sex, and race/ethnicity. A detailed medical history was obtained based on participants’ recall and chart review in addition to a targeted physical exam. A comprehensive COVID-19-specific history was also obtained, including date of symptom onset, disease-related symptoms, date of first positive COVID-19 PCR test, hospitalization status, and COVID-19 therapy received. Physical function was assessed using a work productivity questionnaire and a 6-minute walk test. Initial results involving disease-related symptoms and physical function have been published previously^23^. Finally, blood was drawn to obtain plasma and peripheral blood mononuclear cells (PBMCs), which have been biobanked for inflammatory cytokine assessment and immune profiling.

### Psychiatric Symptom Assessments

During the initial study visit, psychiatric symptoms and quality of life were captured through administration of a battery of self-report questionnaires, including the Patient Health Questionnaire-9 (PHQ-9)^24^ to measure depression, the Generalized Anxiety Disorder 7-item (GAD-7)^25^ to assess anxiety, and the 36-Item Short-Form Health Survey (SF-36)^26^ to measure quality of life. Clinically significant psychiatric symptoms were defined as PHQ-9 and/or GAD-7 ≥ 10 corresponding to a least moderate depression and/or anxiety.

### Neurocognitive Assessments

A battery of neuropsychological assessments was also administered at the first study visit to assess neurocognitive functioning across several key domains, including sustained attention, working memory, processing speed, executive function, selective attention, and recall memory. Assessments were normed to a wide variety of clinical presentations and demographic characteristics^27^, which allows us to derive insights about assessment results in the absence of a baseline measurement (i.e., measurement prior to contracting COVID-19) and in the absence of a control group. The following assessments were administered:

#### Continuous Performance Test (sustained attention)

Participants are presented with letters, one by one, and asked to press when the same letter appears twice in a row. This assesses the participant’s ability to sustain attention over an extended period of time, as well as the ability to update information held in short-term stores of working memory. A normalized score for reaction time was computed for each subject on this task and used for subsequent analysis.

#### Digit Span (working memory)

Participants are presented with a series of digits on the computer screen (500-ms presentation) and are asked to enter the digits presented. In part 1, participants are required to recall the digits in forward order. In part 2, they are required to recall them in reverse order. In each part, the number of digits in each sequence gradually increases from 3 to 7, with two sequences at each level. Maximum span indicates the maximum number of digits the participant recalled without error. The maximum forward recall span was normed and used as a metric for analyses involving working memory.

#### Choice Reaction Time (processing speed)

To assess processing speed, the participant is asked to press the illuminated circle as quickly as possible using the left and right arrow keys on the keyboard. Reaction time was recorded from this task and normalized as a measurement of performance.

#### Maze (executive function)

Participants are asked to discover the hidden path through a grid of circles (by trial and error) presented on the computer screen. This task measures how quickly the participant learned the route through the maze and their ability to remember that route, thus assessing executive function abilities. A normalized score for completion time was chosen as the metric to represent performance for this task in analyses.

#### Verbal Interference Stroop (selective attention)

Participants are presented with colored words (red, yellow, green, or blue), one at a time. They are asked to identify the name of each colored word (ignoring the color of the word), then identify the color of each word (ignoring the name of the color) within 30 seconds. Accuracy for identifying the color of each word was used as a representation of selective attention in subsequent analyses.

#### Verbal Learning and Memory (recall memory)

Participants are presented with a list of 20 words with one word presented at a time. They are then presented with 3 word options from which they must select the recalled word. There are a total of 3 trials. The sum of the immediate recall accuracy for all 3 trials was normed and used as a scoring metric for this task.

### Statistical Analyses for Demographic, Psychiatric, and Neurocognitive Variables

Statistical analyses were run in the python 3 environment and R. Groups were compared using independent samples t-tests, chi-squared tests, and Mann-Whitney U rank tests depending on the scale and distribution of the variables. Scoring for the SF-36 was standardized using US population averages^28^. Webneuro scores were standardized to a healthy reference population matched to our sample by age, biological sex, and education. The signs for scores of reaction and completion times of Webneuro tests were reversed such that all scores above zero represented performance better than average and scores below zero represented performance that was lower than average. Means were then calculated for each of the measures of interest, selected from the six neuropsychological tasks, and one sample t-tests were run comparing each mean to zero. FDR correction using the Benjamini-Hochberg Procedure was implemented on resultant p-values. k-means clustering using minimization of the sum of squared Euclidean distances was applied to the neurocognitive data to separate participants into groups. The number of clusters was determined using the elbow method.

#### Missingness

Data for PHQ-9 was prorated for one subject who had a missing item to bring the sample size to n = 100 by replacing the missing item with the mean of the remaining items. Missing values for the SF-36 and WebNeuro measurements in participants who completed at least one item were replaced with the median score of the group for each missing variable in order to achieve a sample size of n = 96 and n= 89, respectively.

### Structural MRI

In a subset of participants (n=15), a T1-weighted anatomical MRI image was collected after the self-report questionnaires and neurocognitive variables and reviewed by a neuroradiologist to detect any structural olfactory abnormalities. Neuroimaging was completed at the Center for Neurobiological Imaging (CNI) at Stanford University using a GE Discovery MR750 scanner. The parameters for the anatomical T1 image were as follows: TE = 3.828 ms, TR = 3s, FA = 8, acquisition time = 8:33, field of view = 256 × 256 mm, 3D matrix size = 320 × 320 × 230, slice orientation = sagittal, angulation to AC-PC line, receiver bandwidth = 31.25 kHz, fat suppression = no, motion correction = PROMO, voxel size = 0.8 mm isotropic.

### Functional MRI

In the same subset that completed structural MRI (n = 15), a series of functional MRI (fMRI) assessments were run to engage brain circuits that may show dysfunction in post-COVID-19 patients. The parameters were as follows: TE = 27.50 ms, TR = 2 s, FA = 77, acquisition time = 5:08 (Go-NoGo and Viewing of Facial Emotion), field of view = 222 × 222 mm, 3D matrix size = 74 × 74 × 45, slice orientation = axial, angulation to AC-PC line, phase encoding = PA, number of volumes = 151, calibration volumes = 3, voxel size = 3 mm isotropic. The fMRI assessments included:

#### Go-NoGo Task

The Go-NoGo Task has been established as a robust probe of the cognitive control circuit. It was used to assess response inhibition by contrasting ‘NoGo’ responses vs. ‘Go’ responses. 180 Go and 60 NoGo stimuli are presented in pseudorandom order; 500 ms each with an interstimulus interval of 750 ms^29^. In the ‘Go’ trials, participants are required to press on GREEN stimuli (the word ‘press’), while in the ‘NoGo’ trials, participants are required to withhold presses on RED stimuli (**Fig. 2**).

**Fig. 2.**
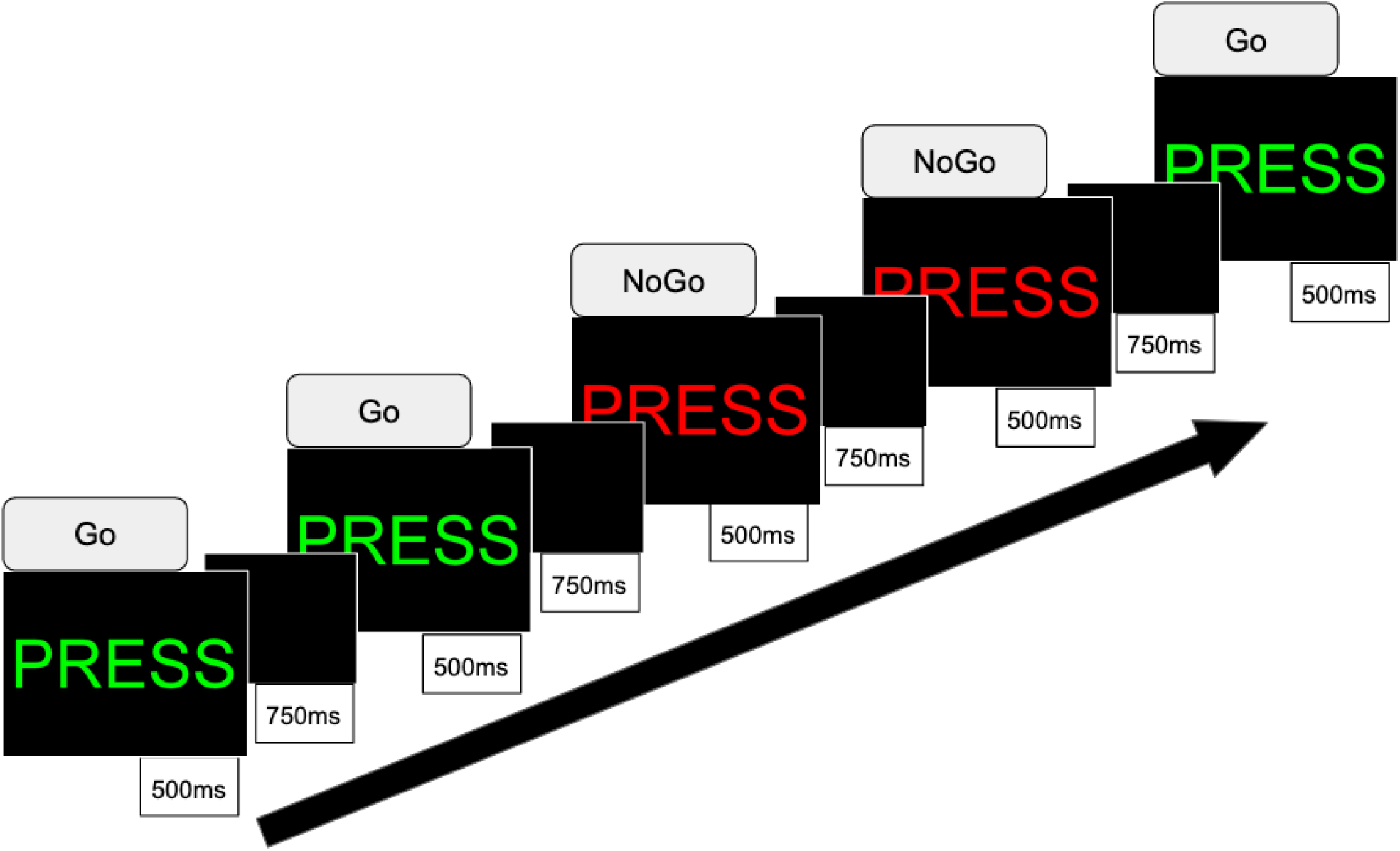
Go-NoGo Task

#### Viewing of Facial Emotion Task

Participants consciously view facial expressions of emotion to probe automatic bottom-up activation of the positive and negative affect circuits^30^. Stimuli are from a standardized series of facial expressions of reward-related emotions (happy), threat-related emotions (fear, anger), and loss related emotions (sad), along with neutral expressions. Stimuli were modified such that the eyes are presented in the central position of the image. Participants are instructed to actively attend to the faces stimuli in order to answer post-scan questions about these faces, and we control for active attention by monitoring alertness with an eye tracking system. To examine the positive affect circuit, happy faces are contrasted with neutral, while for the negative affect circuit, we examine threatening vs. neutral faces and sad vs. neutral faces. Each face is presented for 500 ms with an interstimulus interval of 750 ms.

### Pre-processing, Selecting Regions of Interest, and Statistical Analyses

Pre-processing and data analyses were performed using Statistical Parametric Mapping (SPM) software (Wellcome Department of Cognitive Neurology) implemented in MATLAB and FSL^31^ following previously established procedures^32, 33^. Briefly, pre-processing of functional data included realignment and unwarping, normalization to a standardized template, and smoothing. Quality control diagnostics included removing scans with scanner artefacts and signal dropout. Our QC also utilized standardized pipelines of MRIQC^34^ and fMRIPrep^35^ to measure head motion and for modality-specific quality assessments.

After removing task effects, residual time courses from three fMRI tasks were used for intrinsic functional connectivity (FC) estimation for key nodes in the attention circuit, default mode network (DMN), and the salience circuit. The same data was also used to quantify seed-to-voxel FC seeded in the ventromedial prefrontal cortex (vmPFC) given evidence that this region acts as a central hub for reduced FC associated with increased inflammation^13^. We selected regions of interest (ROIs) for the three functional networks and the vmPFC based on a prior synthesis of the literature^36^ and quantified them using an established systematic procedure. Masks to define these *a priori* regions were generated using the meta-analytic platform Neurosynth^37^ with peaks identified using the Analysis of Functional Neuroimages (AFNI) 3dExtrema function. We imposed a restriction that each peak has a minimum z-score of 6 and each region extends no farther than 10mm from the peak.

For the cognitive control and negative and positive affect circuits, we examined brain-wide activation as elicited by the Go-NoGo and Viewing of Facial Emotion Tasks. We ran one sample t-tests comparing the mean of the IRIS Neurostudy participants standardized to a healthy control sample (n=50), which we independently collected as part of another study^38^, to a mean of zero. We report results at the voxel-level p < 0.001 (uncorrected) and cluster level p < 0.05 (FWE-corrected) for seed-based FC and activation results, and at the edge level p < 0.01 (uncorrected) and component level p < 0.05 (FWE-corrected) for pairwise FC results via the network-based statistic.

Because, to our knowledge, dysfunction as assessed by fMRI has not been previously characterized in post-COVID-19 patients and in order to suggest regions of focus for future studies, we also report results significant at the liberal threshold of voxel-level p < 0.1 (uncorrected) corresponding to a z-score > 1.64. Additionally, we show FC and activation maps for each subject standardized to the healthy control sample and thresholded at z > 1 in supplemental figures.

## Results

### Demographic Characteristics

Demographic characteristics and psychiatric symptoms for the full sample and divided by hospitalization status can be found in **Table 1**. The sample is comprised of 52 females and 47 males with a mean age 44.9 ± 14.3 and age range of 20-83 years. Of the 99 participants with hospitalization status data, 29 were hospitalized (29.3%) and 12 (12.1%) were admitted to the ICU. Hospitalized participants were significantly older (p=0.008) without significant differences in other demographic characteristics.

**Table 1.** Demographic characteristics, psychiatric symptoms, and quality of life

| Variable | All | Non-hospitalized | Hospitalized | p-value |
| --- | --- | --- | --- | --- |
| **Age (Mean, SD)** | **n=99** | **n=70** | **n=29** |  |
|  | 44.9 (14.3) | 42.2 (13) | 51.3 (15.3) | **0.008** |
| **Sex (n, %)** | **n=99** | **n=70** | **n=29** |  |
| Male | 47 (47.5%) | 30 (42.9%) | 17 (58.6%) | ns |
| Female | 52 (52.5%) | 40 (57.1%) | 12 (41.4%) |  |
| **Race/Ethnicity (n, %)** | **n=100** | **n=71** | **n=29** |  |
| Asian | 19 (19%) | 9 (12.7%) | 10 (34.5%) | ns |
| Pacific Islander | 1 (1%) | 1 (1.4%) | 0 (0%) |  |
| Black or African American | 2 (2%) | 1 (1.4%) | 1 (3.5%) |  |
| White | 76 (76%) | 59 (83.1%) | 17 (58.6%) |  |
| Other | 2 (2%) | 1 (1.4%) | 1 (3.5%) |  |
| **Depression Severity (n, %)** | **n=100** | **n=71** | **n=29** |  |
| Minimal/Mild Depression | 83 (83%) | 63 (88.7%) | 20 (69%) | **0.04** |
| Moderate/Severe Depression | 17 (17%) | 8 (11.3%) | 9 (31%) |  |
| **Anxiety Severity (n, %)** | **n=99** | **n=71** | **n=28** |  |
| Minimal/Mild Anxiety | 86 (86.9%) | 61 (85.9%) | 25 (89.3%) | ns |
| Moderate/Severe Anxiety | 13 (13.1%) | 10 (14.1%) | 3 (10.7%) |  |
| **Quality of Life (Standardized Mean, SD)** | **n=96** | **n=67** | **n=29** |  |
| Physical functioning | -0.08 (1.11) | 0.12 (0.95) | -0.53 (1.32) | **0.03** |
| Role functioning/physical | -0.36 (0.92) | -0.30 (0.95) | -0.48 (0.85) | ns |
| Role functioning/emotional | -0.16 (0.87) | -0.15 (0.93) | -0.19 (0.75) | ns |
| Energy/fatigue | -0.38 (0.83) | -0.38 (0.83) | -0.38 (0.85) | ns |
| Emotional well-being | -0.51 (0.88) | -0.51 (0.90) | -0.49 (0.83) | ns |
| Social functioning | -0.54 (1.27) | -0.52 (1.25) | -0.58 (1.35) | ns |
| Pain | -0.07 (1.05) | 0.04 (1.06) | -0.32 (1.01) | ns |
| General health | -0.46 (1.04) | -0.41 (0.97) | -0.59 (1.23) | ns |

Depression and anxiety were measured using the Patient Health Questionnaire-9 (PHQ-9) and the Generalized Anxiety Disorder 7-item (GAD-7), respectively. Minimal/mild was defined as < 10 and moderate/severe as ≥ 10 on both scales. The 36-Item Short-Form Health Survey (SF-36) was used to assess quality of life.

### Psychiatric Symptoms

Over 1/5 of participants (22%) had moderate to severe depression and/or anxiety symptoms, indicative of clinically significant psychiatric impairment (**Fig. 3A**). A more detailed breakdown of depression/anxiety symptoms revealed that 17% of the IRIS Neurostudy cohort reported moderate to severe depression (**Fig. 3B**), while 13.1% had symptoms consistent with moderate to severe anxiety (**Fig. 3C**). Hospitalized participants were more likely to have clinically significant depression as compared to non-hospitalized participants (**Table 1;** p = 0.04), although anxiety did not differ. Quality of life measures did not differ significantly between hospitalized and non-hospitalized participants except in the domain of physical functioning, which was worse in hospitalized participants (**Table 1;** p = 0.03).

**Fig. 3.**
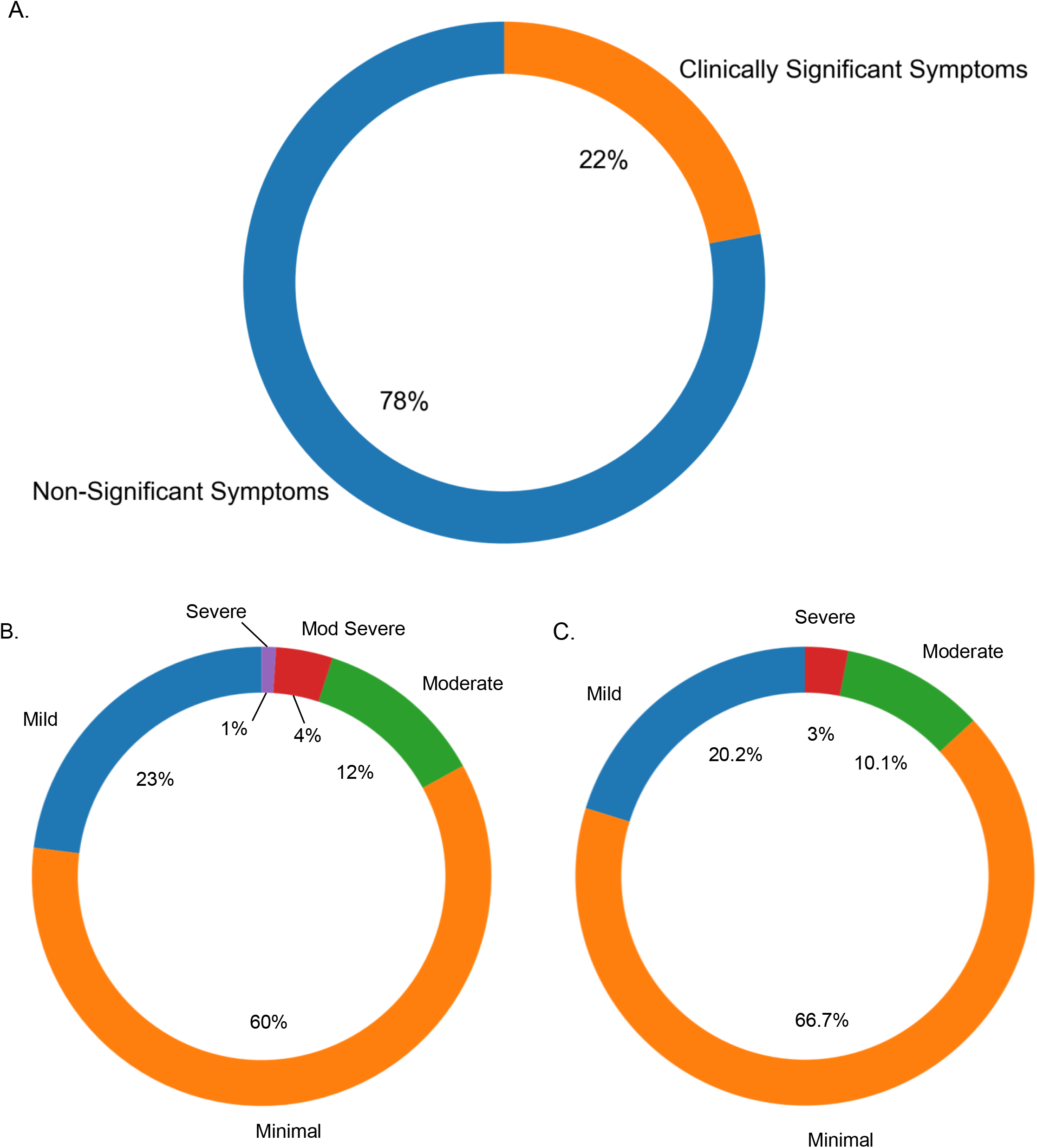
**(A)** Proportion of participants for whom depression (PHQ-9) and/or anxiety (GAD-7) was at least in the moderate range, indicative of clinically significant psychiatric symptoms. **(B)** Breakdown of scoring on the PHQ-9 and (**C**) on GAD-7 based on reported symptom severity.

### Neurocognitive Impairments

Participants showed a profile consistent with ‘brain fog’ characterized by significant impairment in the domains of sustained attention, working memory, processing speed, and executive function when compared to standardized age, sex, and education matched healthy reference norms (adjusted p’s ≤ 0.001, **Fig. 4**; **Table 2**). k-means clustering applied to the neurocognitive data yielded two clusters (**Fig. 5A**). One cluster, 41.6% of the cohort, was ‘impaired’ across all domains; the other was relatively ‘intact’ with the exception of sustained attention, which was impaired in both clusters (**Fig. 5B)**. There were no statistically significant differences between measures of psychiatric symptoms (**Fig. 5C**) when comparing the two clusters.

**Table 2.** Mean neurocognitive performance

| Neurocognitive domain | Test | Standardized mean | Adjusted p |
| --- | --- | --- | --- |
| Sustained Attention | Continuous Performance | -1.21 | **3.45 x 10<sup>-15</sup>** |
| Working Memory | Digit Span | -0.721 | **0.00001** |
| Processing Speed | Trails B | -0.301 | **0.001** |
| Executive Function | Maze | -0.443 | **0.0005** |
| Selective Attention | Stroop | -0.220 | ns |
| Recall Memory | Verbal Learning and Memory | -0.008 | ns |

**Fig. 4.**
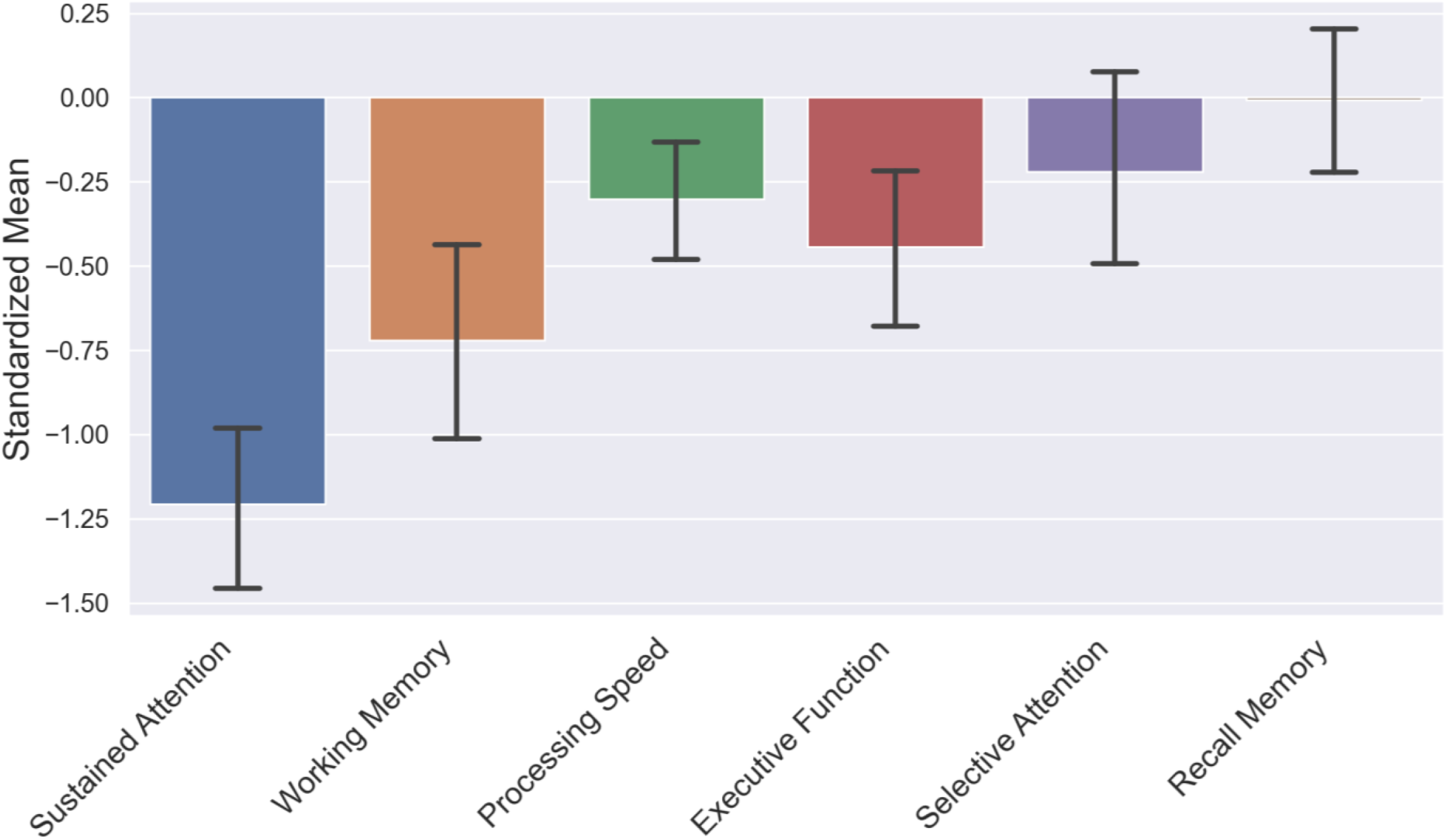
Standardized mean performance for 6 neurocognitive variables. Error bars represent the 95% confidence interval.

**Fig. 5.**
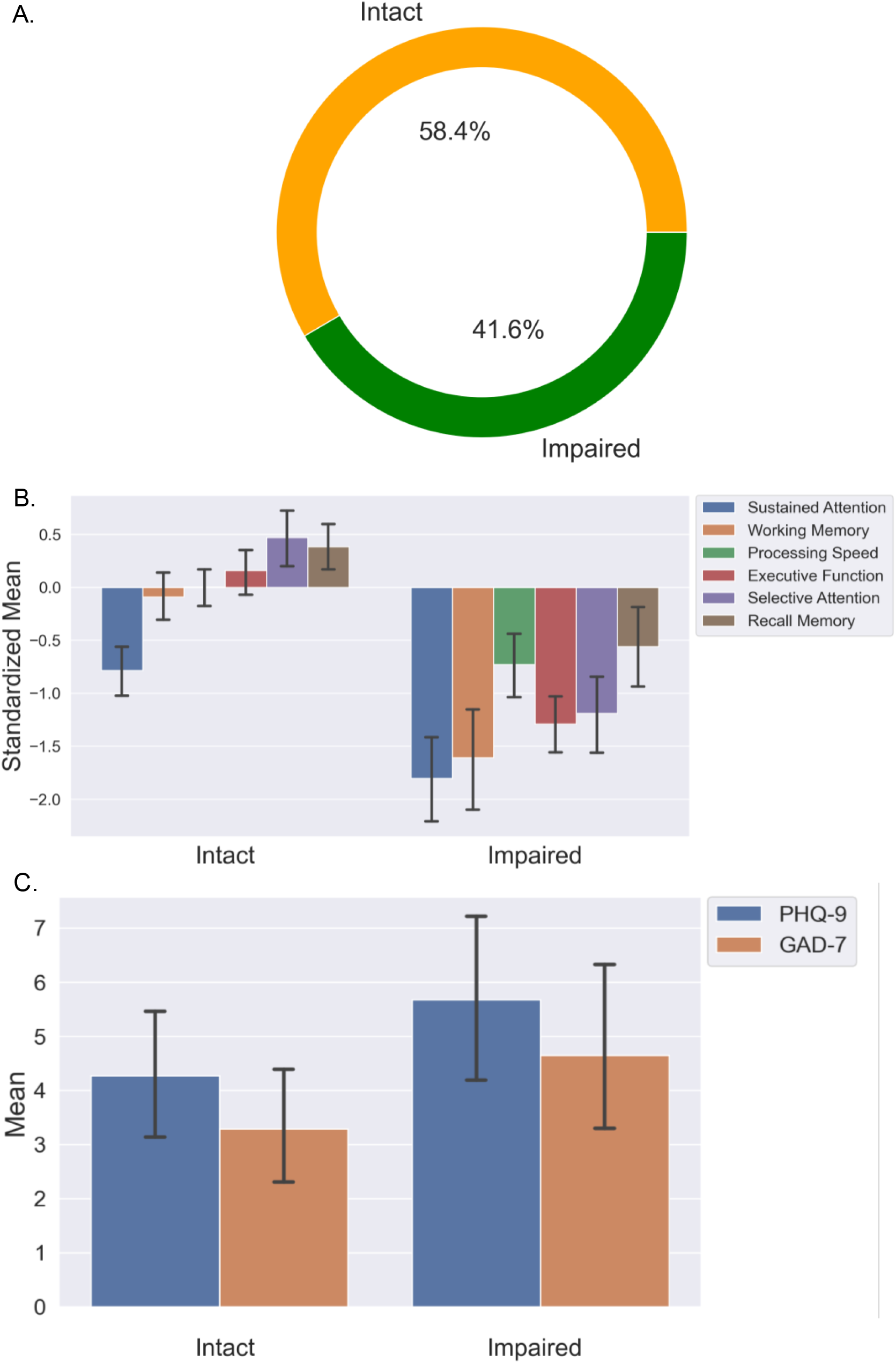
(**A**) Proportion of participants who display ‘impaired’ versus relatively ‘intact’ cognition defined using clustering analysis. (**B**) Standardized mean neurocognitive performance within each cluster. Error bars represent the 95% confidence interval. (**C**) Comparison of psychiatric symptoms (PHQ-9 & GAD-7) between cognition clusters. Error bars represent the 95% confidence interval.

### Olfactory Bulb Abnormalities Assessed with Structural MRI

Clinical reads of anatomical MRI scans were completed for the 15 participants who underwent MRI by a trained neuroradiologist, and eight participants (53.3%) were found to have small olfactory bulbs (OB). All eight of these participants reported new onset anosmia during their first study visit. OB volumes and sulcus depths were also quantified (**Table 3)**. The minimum normative value for OB volume has previously been defined as 58 mm^3^ in individuals younger than 45 years old and 46 mm^3^ in individuals older than 45 years old^39^. According to this metric, two additional participants qualified as having small olfactory bulbs on the right, noting that only 13 participants could be quantified due to degraded image quality for two of them. Using the minimum normative value for OS depth as 7.5 mm^22^, 11 out of 13 participants (84.6%) with measurements met criteria for shallow OS. We did not find any significant relationships between OB volume or OS depth and clinically significant psychiatric symptoms or membership in the cognitively impaired cluster. **Fig. 6** shows representative T1-weighted images of small and normal OB volumes.

**Table 3.** Olfactory findings in T1-weighted MRI

| ID | Age Range | Anatomical finding | Left OB volume (mm<sup>3</sup>) | Right OB volume (mm<sup>3</sup>) | Left OS depth (mm) | Right OS depth (mm) | New Onset Anosmia |
| --- | --- | --- | --- | --- | --- | --- | --- |
| IRIS008 | 36 - 41 | Normal olfactory bulb | Image quality degraded by motion artifact |  |  |  | Not present |
| IRIS010 | 45 - 50 | Normal olfactory bulb | 71.13 | 67.23 | 6.74 | 6.85 | Present |
| IRIS011 | 36 - 41 | Small olfactory bulb | 40.23 | 39.87 | 5.89 | 6.65 | Present |
| IRIS021 | 60 - 65 | Normal olfactory bulb | 67.40 | 70.60 | 6.14 | 6.48 | Present |
| IRIS029 | 30 - 35 | Small olfactory bulb | 48.90 | 44.20 | 6.12 | 5.33 | Present |
| IRIS032 | 30 - 35 | Small olfactory bulb | 44.87 | 44.00 | 6.71 | 6.60 | Present |
| IRIS035 | 45 - 50 | Small olfactory bulb | 42.33 | 42.23 | 6.97 | 6.80 | Present |
| IRIS038 | 36 - 41 | Small olfactory bulb | 45.30 | 37.90 | 5.67 | 6.02 | Present |
| IRIS040 | 45 - 50 | Small olfactory bulb | 37.40 | 41.03 | 7.48 | 7.51 | Present |
| IRIS043 | 24 - 29 | Small olfactory bulb | 45.20 | 42.77 | 7.43 | 6.80 | Present |
| IRIS044 | 24 - 29 | Normal olfactory bulb | 60.57 | 59.10 | 7.68 | 7.91 | Present |
| IRIS045 | 24 - 29 | Normal olfactory bulb | 55.90 | 57.83 | 5.64 | 6.11 | Present |
| IRIS048 | 24 - 29 | Normal olfactory bulb | 62.97 | 57.70 | 7.74 | 7.28 | Present |
| IRIS084 | 51 - 55 | Normal olfactory bulb | Image quality degraded by motion artifact |  |  |  | Not present |
| IRIS085 | 36 - 41 | Small olfactory bulb | 42.93 | 41.73 | 5.52 | 6.06 | Present |

Abbreviations: Olfactory bulb (OB); Olfactory sulcus (OS).

**Fig. 6.**
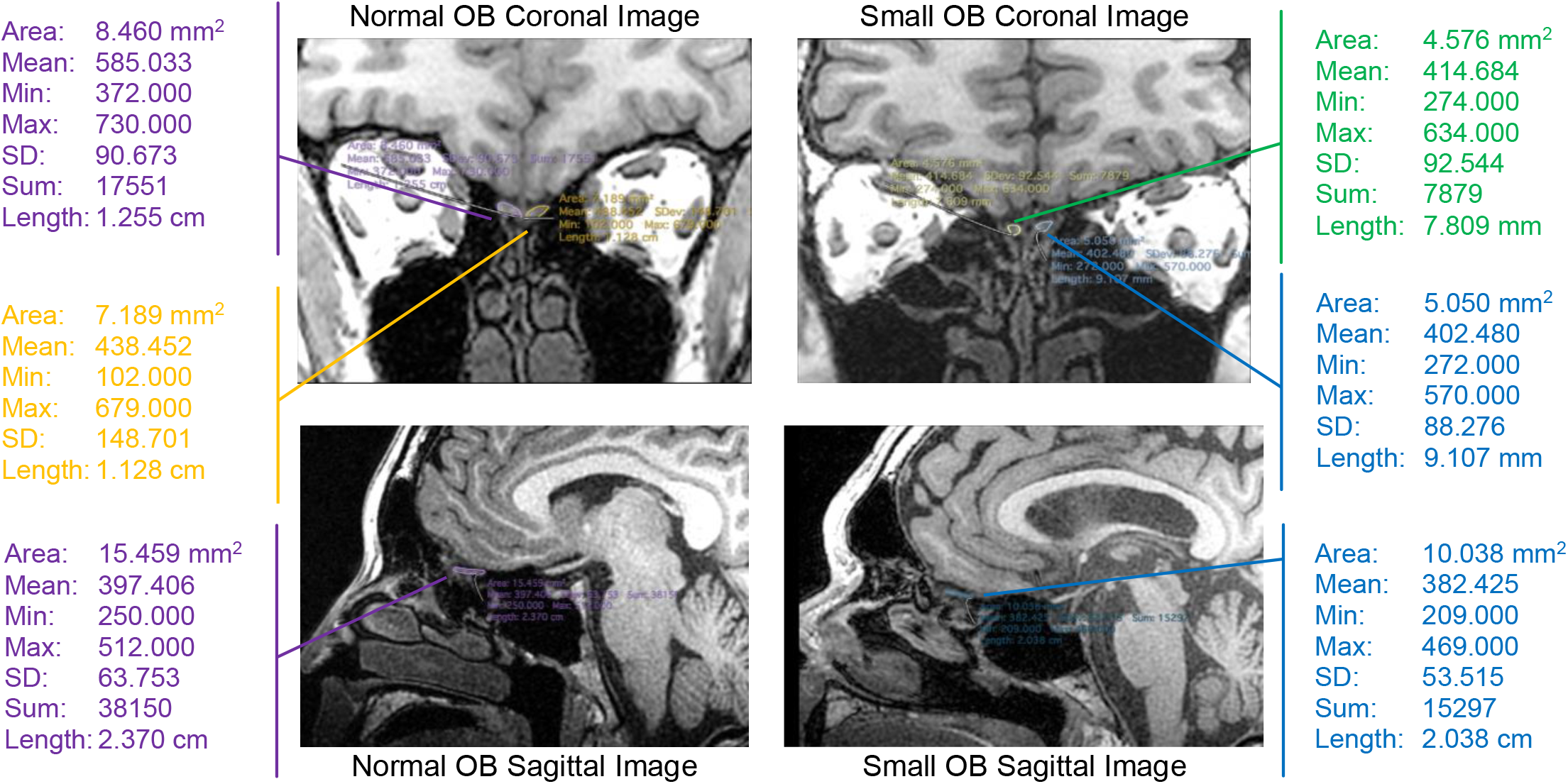
Representative T1-weighted structural images of small (IRIS035) and normal (IRIS010) olfactory bulb volumes. Abbreviations: Olfactory bulb (OB).

### Neural Circuit Dysfunction Assessed with Functional Magnetic Resonance Imaging (fMRI)

#### Group Results

When examining results for the neuroimaging data collected thus far (n=15), we found that pairwise FC between the medial superior prefrontal cortex (msPFC) and the bilateral precuneus within the attention network was significantly lower for standardized IRIS participants (edge level p < 0.01 (uncorrected) and component level p < 0.05 (FWE-corrected) (**Fig. 7**). We also showed that, for our seed-to-voxel intrinsic FC analysis with vmPFC as the ROI, standardized IRIS participants showed hypoconnectivity between the vmPFC and its surrounding voxels, middle cingulate cortex, and bilateral ventral striatum (voxel-level p < 0.001 (uncorrected) and cluster level p < 0.05 (FWE-corrected) (**Fig. 10**). No brain-wide activation results survived multiple test correction.

**Fig. 7.**
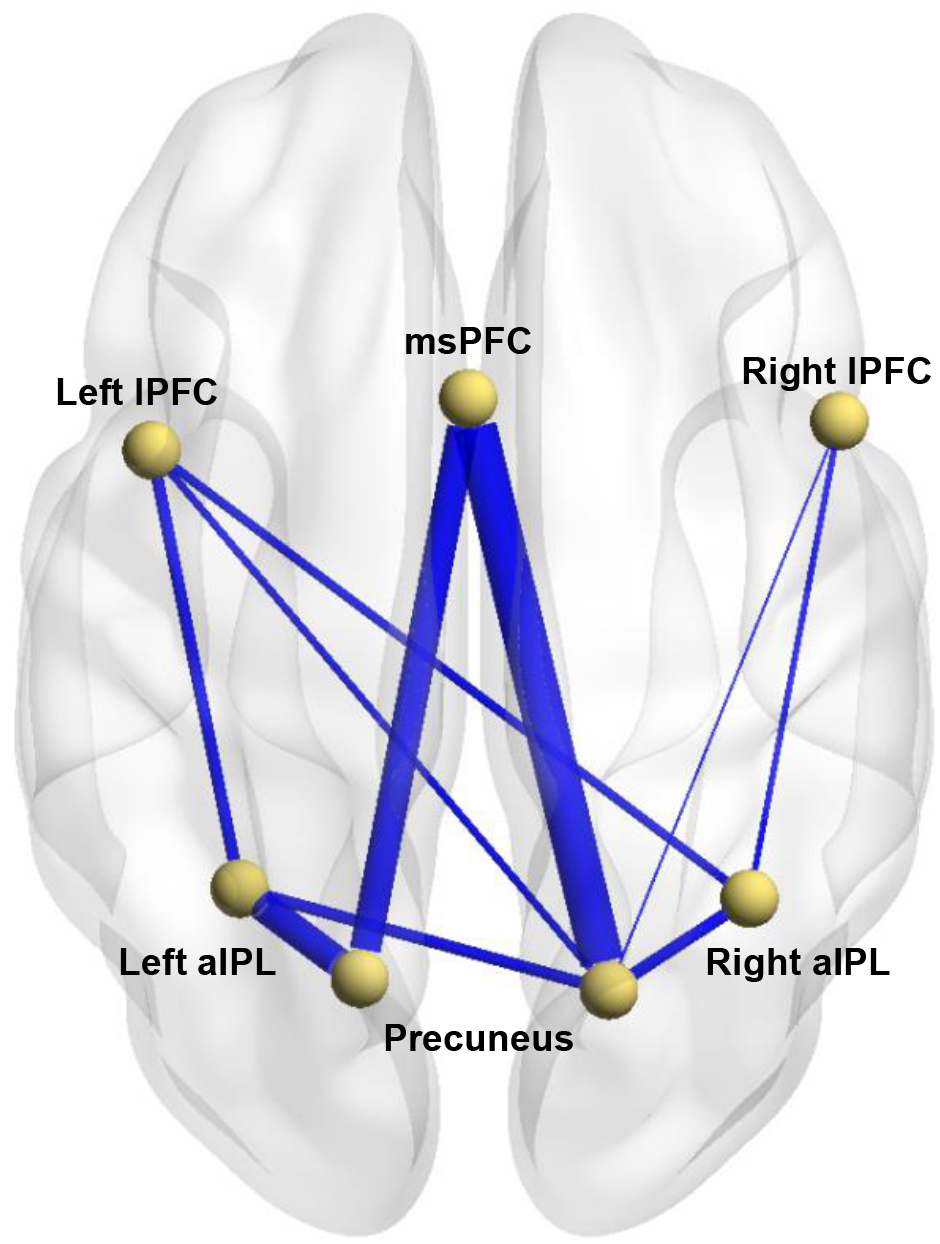
Intrinsic Functional Connectivity Map for the Attention Network (p < 0.1, z > 1.64). Map is standardized to healthy controls with blue indicating lower intrinsic functional connectivity (FC) and red indicating higher FC as compared to controls. The thickness of the lines represents the strength of FC. For this network, we found only lower intrinsic FC at the p < 0.1 level. Abbreviations: aIPL = anterior inferior parietal lobule; lPFC = lateral prefrontal cortex; msPFC = medial superior prefrontal cortex.

At the liberal threshold of voxel-level p < 0.1 (uncorrected), the attention network demonstrated additional nodes with lower pairwise functional connectivity in standardized IRIS participants, including between each lateral prefrontal cortex (lPFC) and the corresponding anterior inferior parietal lobules (aIPL) (**Fig. 7**). Lower pairwise FC was also observed between each aIPL and the corresponding precuneus, the right lPFC and the right precuneus, the left lPFC and the right precuneus, and the left IPFC and the right aIPL. The default mode network showed hyperconnectivity between the right angular gyrus and the anterior medial prefrontal cortex **(Fig. 8**). We did not observe any group differences at the uncorrected p < 0.1 level for the salience network (**Fig. 9**). For our seed-to-voxel intrinsic FC analysis with vmPFC as the ROI, in addition to the result that survived multiple test correction (described above), at the uncorrected p < 0.1 level we also found hypoconnectivity between the vmPFC and dorsolateral prefrontal cortex (DLPFC) as well as hyperconnectivity between the vmPFC and posterior cingulate cortex (PCC) (**Fig. 10**).

**Fig. 8.**
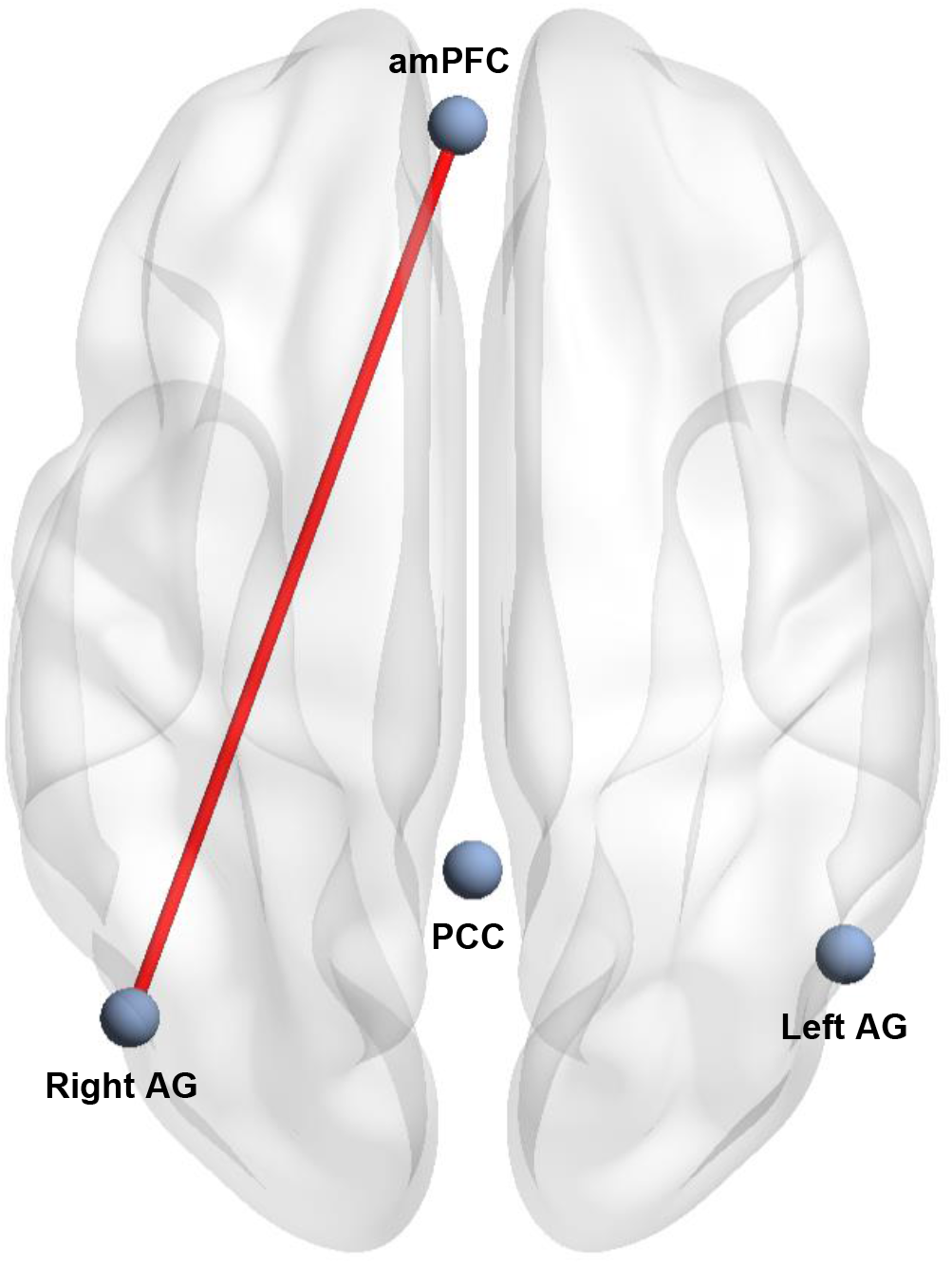
Intrinsic Functional Connectivity Map for the Default Mode Network (p < 0.1, z > 1.64). Map is standardized to healthy controls with blue indicating lower intrinsic functional connectivity (FC) and red indicating higher FC as compared to controls. The thickness of the lines represents the strength of FC. For this network, we found only higher intrinsic FC at the p < 0.1 level. Abbreviations: amPFC = anterior medial prefrontal cortex; AG = angular gyrus; PCC = posterior cingulate cortex.

**Fig. 9.**
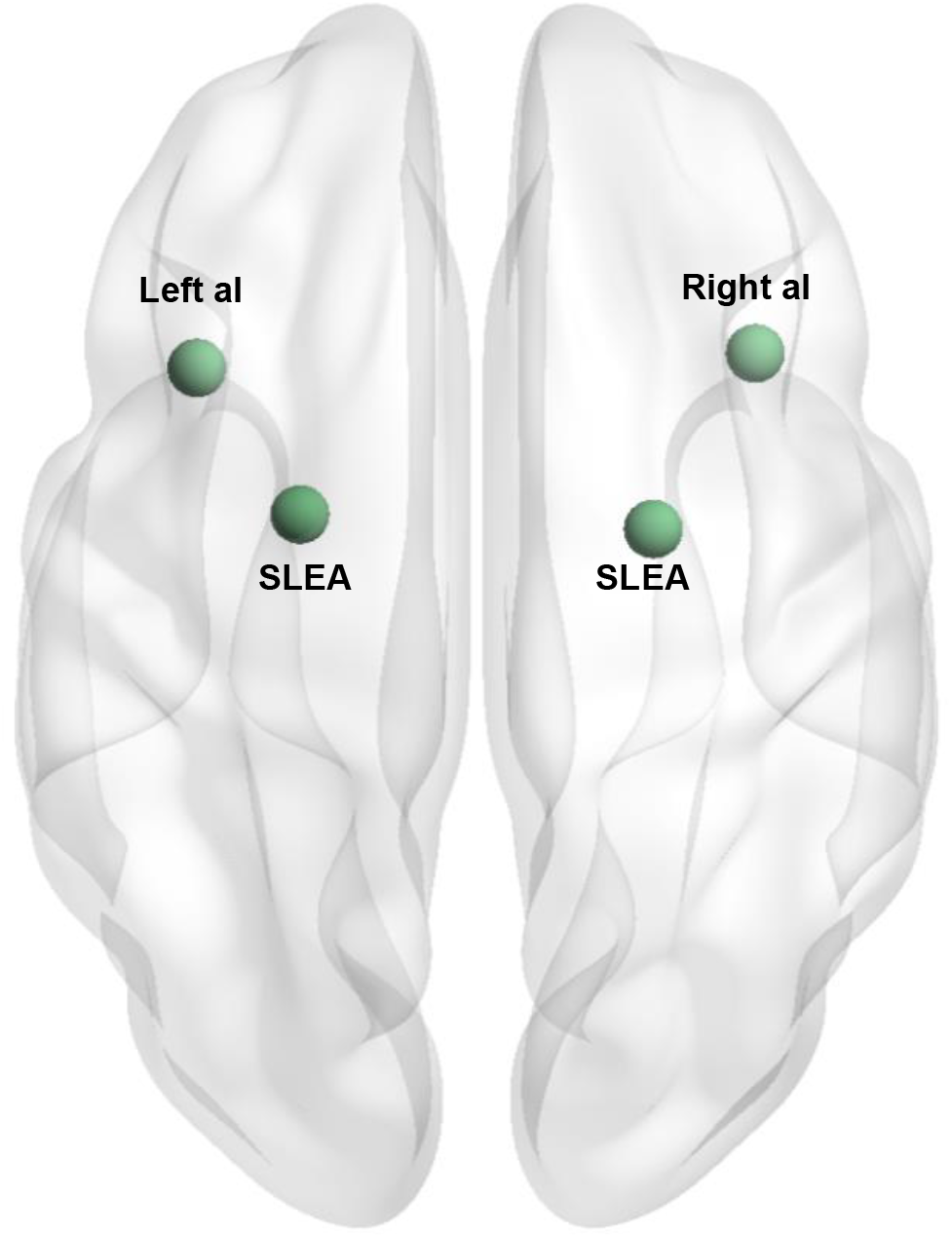
Intrinsic Functional Connectivity Map for the Salience Network (p < 0.1, z > 1.64). Map is standardized to healthy controls with blue indicating lower intrinsic functional connectivity (FC) and red indicating higher FC as compared to controls. The thickness of the lines represents the strength of FC. For this network, we did not find any FC differences at the p < 0.1 level. Abbreviations: aI = anterior insula; SLEA = superior lateral extended amygdala.

**Fig. 10.**
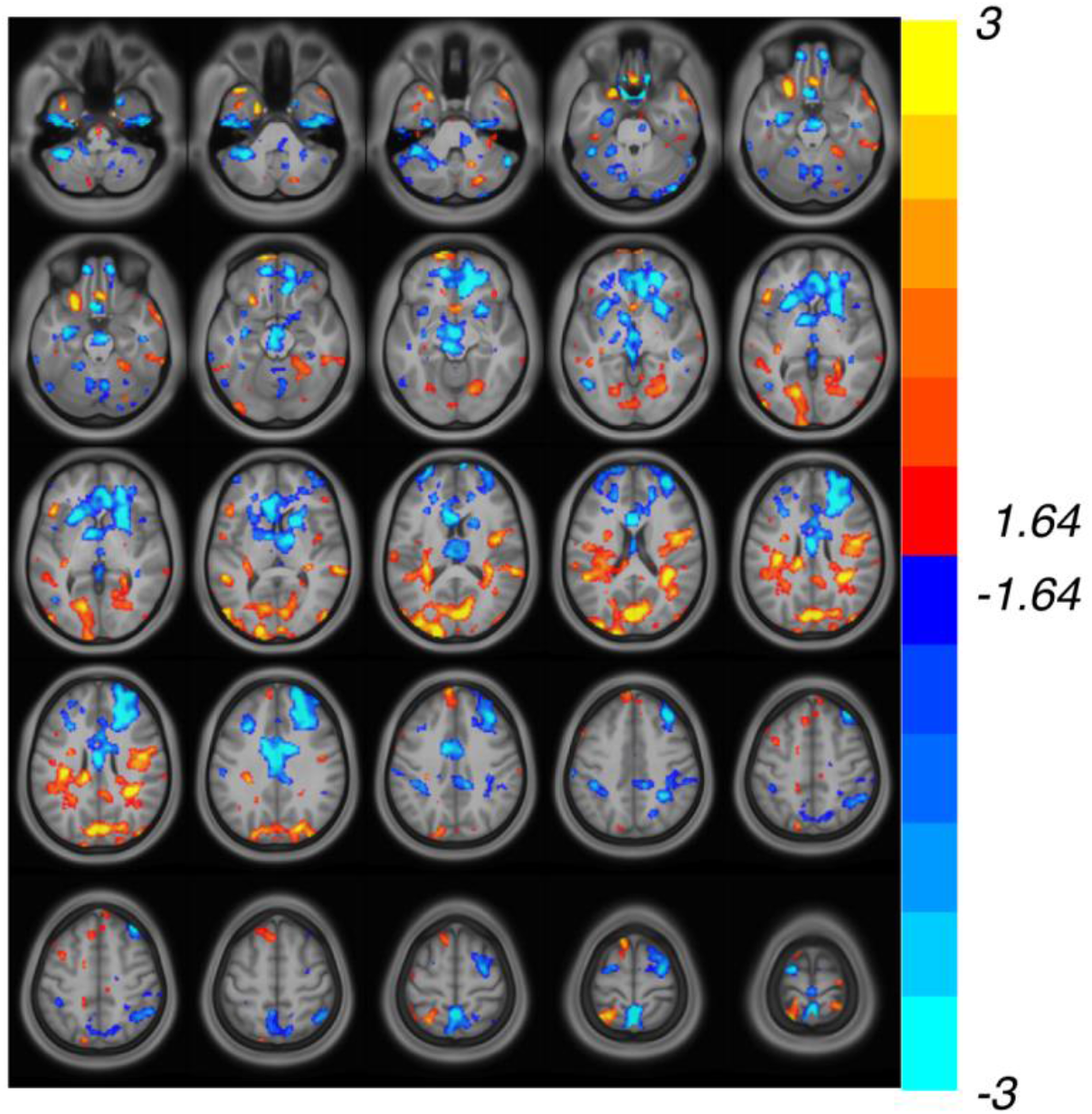
Intrinsic Functional Connectivity Map for Seed in Ventral Medial Prefrontal Cortex (vmPFC) (p < 0.1, z > 1.64). Map is standardized to healthy controls with warm colors representing hyperactivation and cool colors representing hypoactivation.

For brain-wide activation analyses (voxel-level p < 0.001 (uncorrected) and cluster level p < 0.05 (FWE-corrected) with the Go-NoGo task (**Fig. 11**), we observed hypoactivation in the right DLPFC extending to anterior insula, and hyperactivation in the anterior cingulate cortex (ACC), visual cortex, and left temporal gyrus. We also observed a large cluster (voxels = 13732) of hyperactivation that spans multiple regions known to be affected by physiological noise and vessel pulsation. For this reason, we exclude this cluster from our discussion. The happy vs. neutral contrast of the Viewing of Facial Emotion Task revealed hypoactivation in the PCC and mPFC, and hyperactivation of the DLPFC extending to anterior insula, bilateral precentral cortex, and bilateral middle frontal lobe (**Fig. 12**). Finally, for the threatening vs. neutral contrast, we found that the IRIS group showed hyperactivation in the right DLPFC and bilateral middle frontal lobe and hypoactivation in the thalamus and caudate compared to controls (**Fig. 13**), while they showed hypoactivation in the PCC, mPFC, and left superior frontal lobe compared to controls on the sad vs. neutral contrast (**Fig. 14**). Similar to the Go-NoGo Task, there was a large cluster of hypoactivation (voxels = 10147) that spans multiple regions known to be affected by physiological noise and vessel pulsation, and thus, we exclude this cluster from our discussion.

**Fig. 11.**
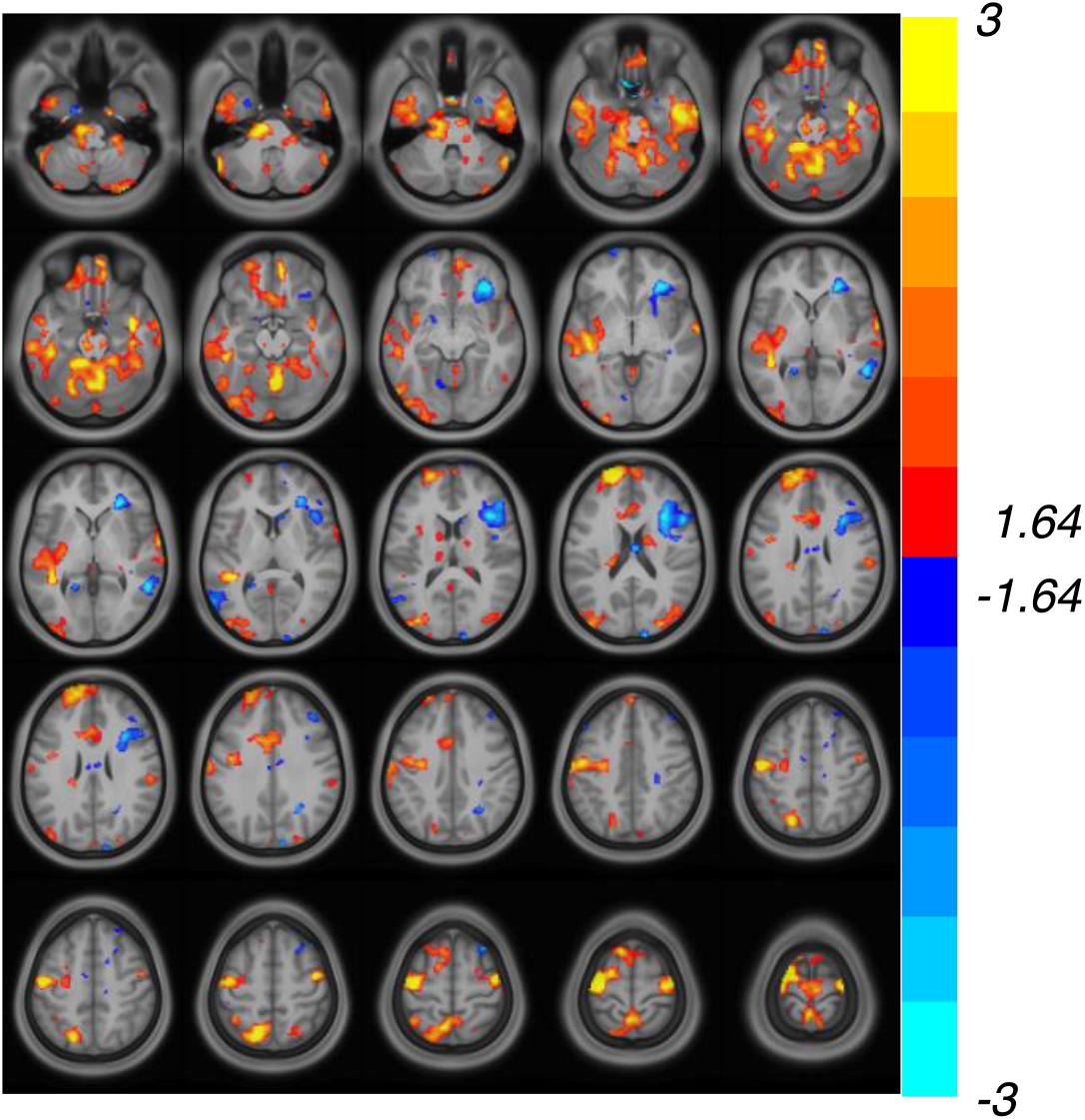
Activation Map for NoGo (cognitive control) Relative to the Go Condition in the GoNoGo Task (p < 0.1, z > 1.64). Map is standardized to healthy controls with warm colors representing hyperactivation and cool colors representing hypoactivation.

**Fig. 12.**
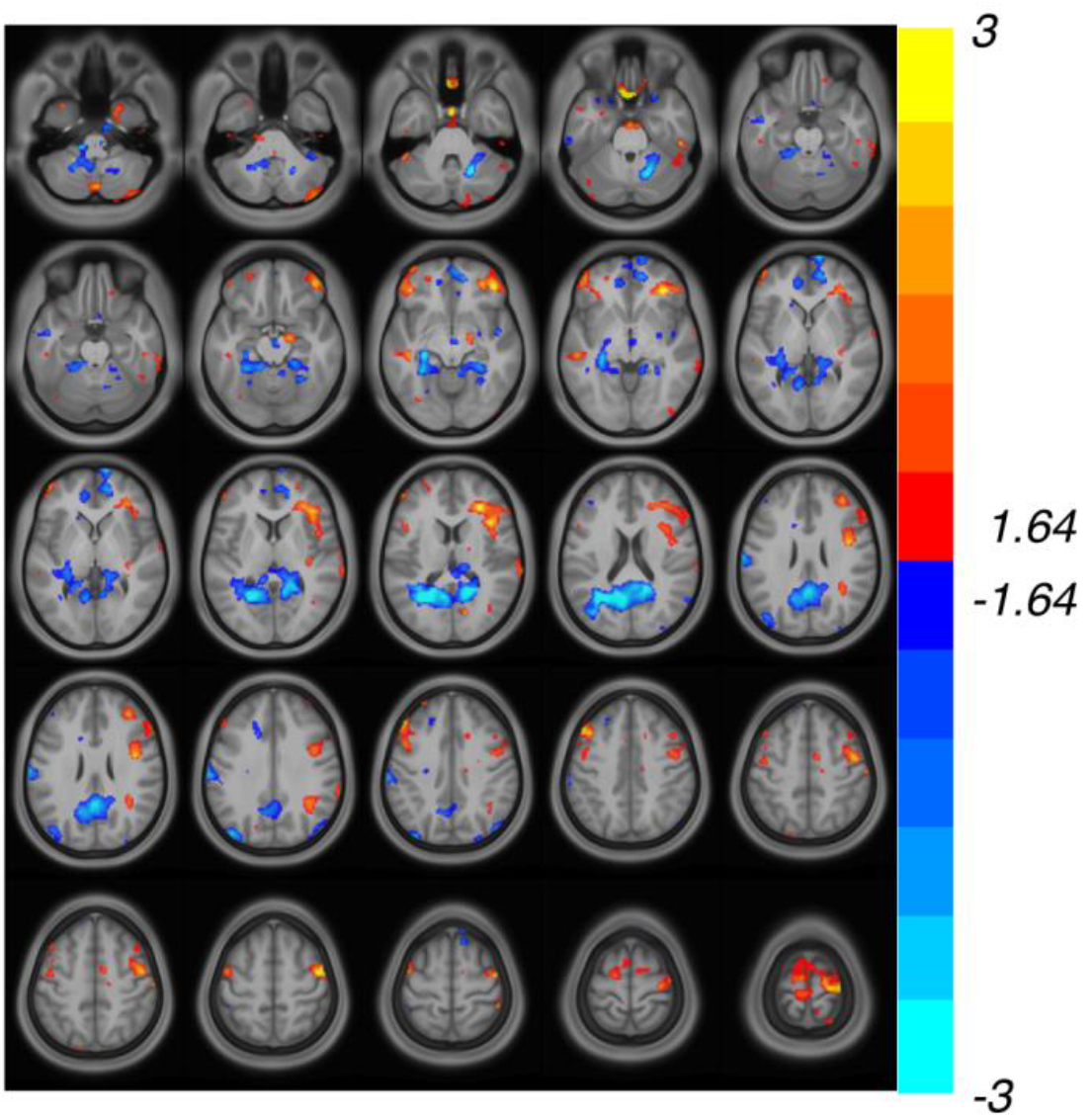
Activation Map for Happy Faces (positive affect) Relative to Neutral in the Viewing of Facial Emotion Task (p < 0.1, z > 1.64). Map is standardized to healthy controls with warm colors representing hyperactivation and cool colors representing hypoactivation.

**Fig. 13.**
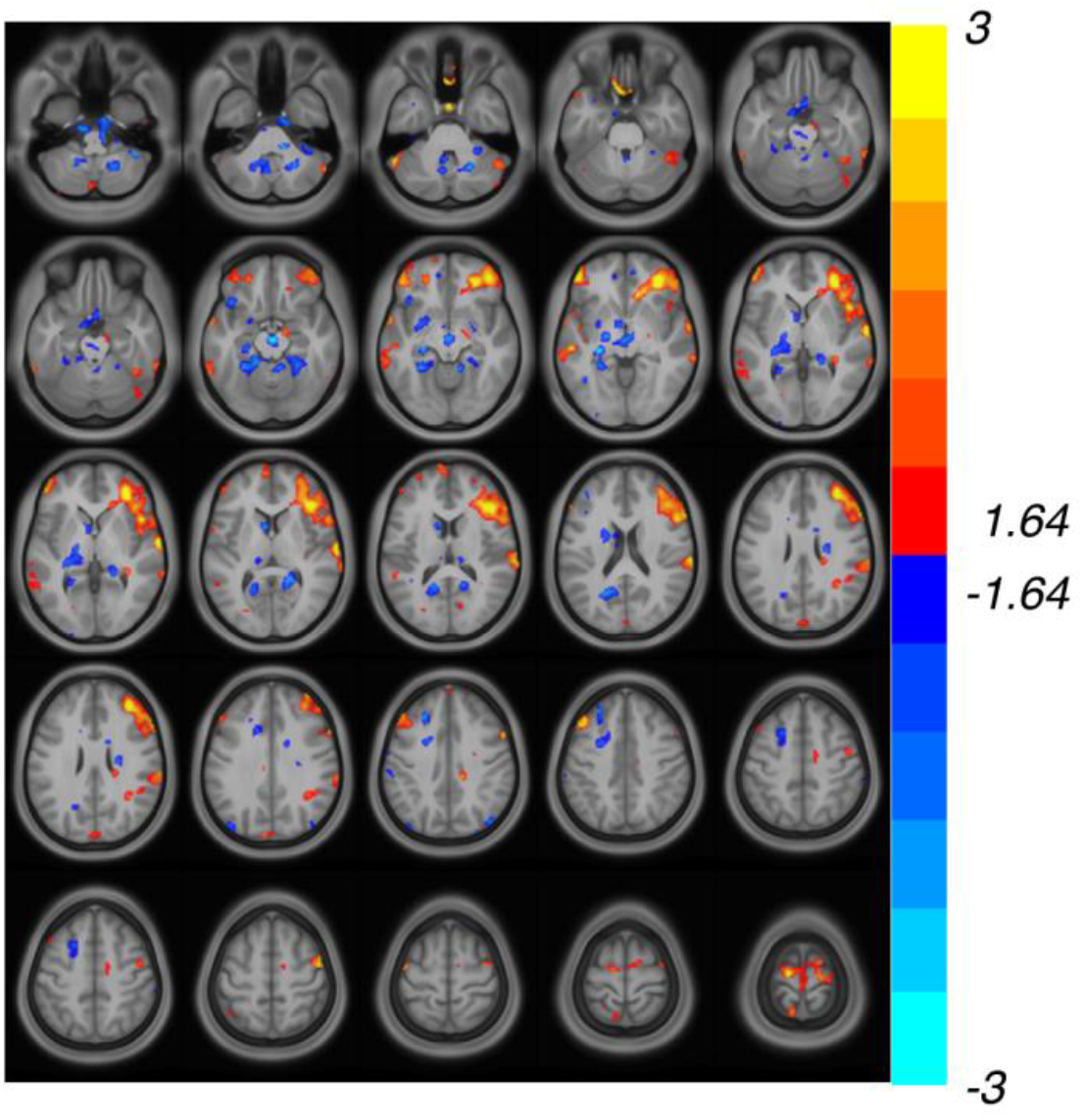
Activation Map for Threatening Faces Relative to Neutral is the Viewing of Facial Emotion Task (p < 0.1, z > 1.64). Map is standardized to healthy controls with warm colors representing hyperactivation and cool colors representing hypoactivation.

**Fig. 14.**
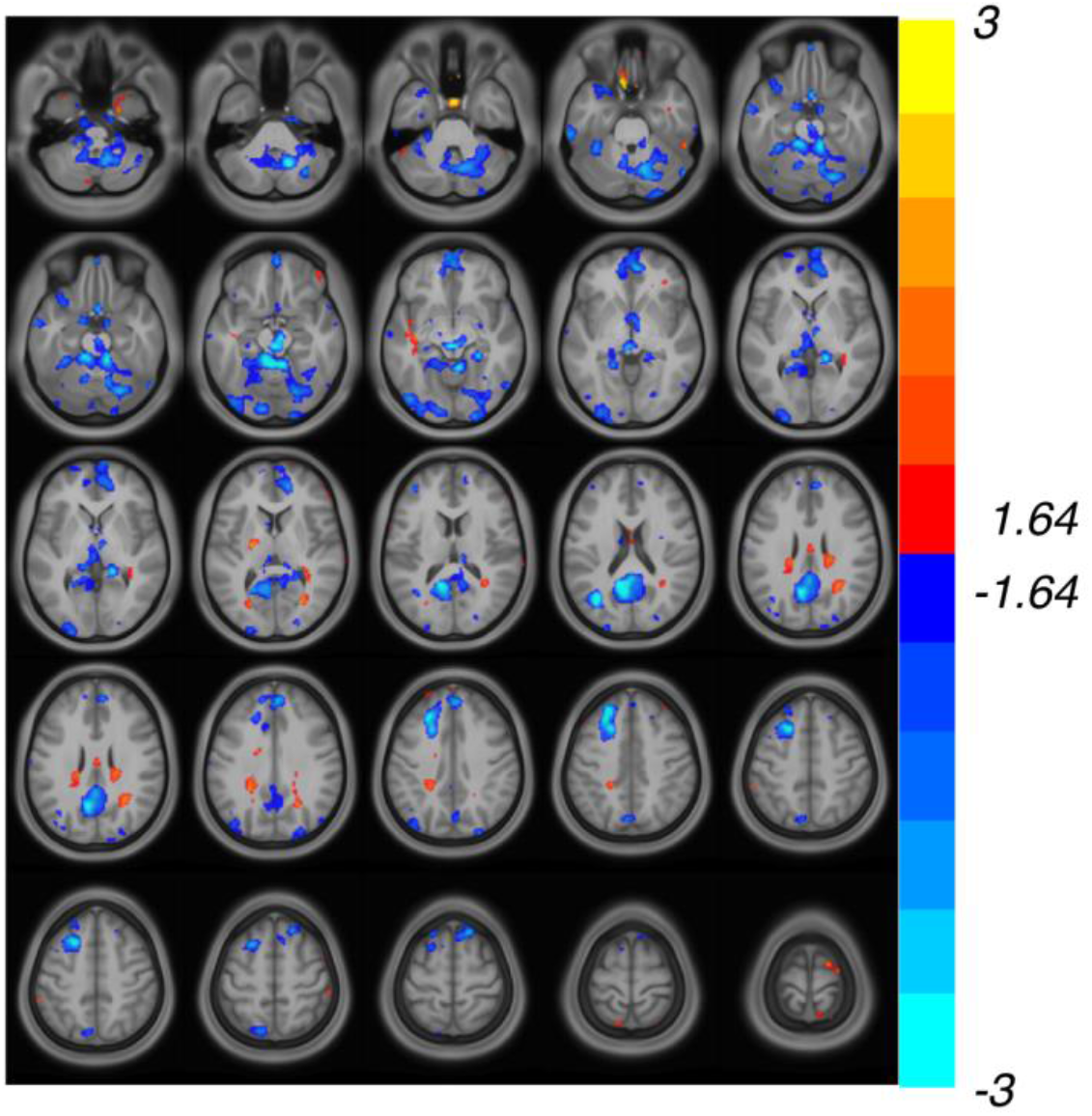
Activation Map for Sad Faces Relative to Neutral in the Viewing of Facial Emotion Task (p < 0.1, z > 1.64). Map is standardized to healthy controls with warm colors representing hyperactivation and cool colors representing hypoactivation.

#### Individual Results

In an attempt to find patterns between the individual pairwise functional connectivity and activation maps and psychiatric symptoms and cognitive performance, we labelled each map with icons indicating whether the participant was in the cognitively impaired cluster and/or had clinically significant psychiatric symptoms on PHQ-9 and/or GAD-7 (**Supplemental Figs. 1-8**). We did not identify any patterns between the maps and the clinical symptoms or cognitive performance.

## Discussion

Collectively, our preliminary results can be broken down and interpreted within four overarching and intersecting domains: psychiatric symptoms, neurocognitive impairments, olfactory anatomical abnormalities, and neural circuit dysfunction.

### Psychiatric Symptoms

Our finding that 22% of the sample had clinically significant psychiatric symptoms in the months following COVID-19 diagnosis aligns with a recent large analysis of electronic medical record data showing that 33.62% of participants diagnosed with COVID-19 met criteria for a psychiatric disorder 6 months later with 12.84% receiving their first such diagnosis^40^. Due to a limited number of participants with structured diagnostic interview results currently, we do not present analyses considering psychiatric diagnoses prior to contracting SARS-CoV-2. However, as we collect this structured interview on additional participants, we will be able to make conclusions from our data about the impact of prior diagnoses on post-COVID-19 psychiatric symptoms. Notably, level of anxiety and impairment in the majority of quality of life domains did not differ significantly between hospitalized and non-hospitalized participants, highlighting that severity of post-COVID-19 neuropsychiatric symptoms and impairments may be independent of infection severity.

### Neurocognitive Impairments

The observed impairment across cognitive domains of sustained attention, working memory, processing speed, and executive function compared with healthy controls is consistent with a profile of ‘brain fog’ post-COVID-19 infection. Our clustering analysis revealed that, while there is a distinction overall between participants within the IRIS Neurostudy cohort who have global cognitive impairment and those who are within the range of healthy cognition, sustained attention remains impaired even in those participants who do not have dysfunction in other domains. This aligns with prior evidence indicating that attention deficits are the most common cognitive dysfunction in post-COVID-19 participants by self-report^15^. Notably, prior work has suggested that impairments in cognition, including attention, in the context of depression respond poorly to conventional antidepressants^41^, suggesting that post-COVID-19 participants with these symptoms may need alternative treatments. Furthermore, there is evidence that elevated levels of peripheral inflammation are correlated with cognitive dysfunction in depression^42^.

### Olfactory Bulb Abnormalities

In our subsample of participants with MRI data, 53.3% of participants were found to have small olfactory bulbs on clinical reads at approximately three months post infection. Of these individuals, all reported new onset anosmia at approximately three months post-infection. Furthermore, OS depths were found to be shallow in 84.6% of participants who had high quality imaging data. These findings align with a previous study showing decreases in both OB volumes (43.5%) and OS depths (60.9%) as compared to normative cut-offs measured one to four months after infection in a sample of COVID-19 participants selected for anosmia^43^. Our results are also consistent with prior case reports, including one showing OB atrophy and anosmia at approximately one month post-infection that persisted at four months^20^ and another showing OB atrophy in association with anosmia in a patient one month after COVID-19 diagnosis in comparison to pre-COVID-19 imaging^21^. The latter study is consistent with a potential causal relationship whereby SARS-CoV-2 may enter the brain through the olfactory epithelium, damaging the olfactory bulb and causing an impaired sense of smell. Indeed, while the evidence is mixed, there are some animal and human studies suggesting that SARS-CoV-2 is capable of viral neuroinvasion via the olfactory nerve or other routes^17^, similar to HSV1, varicella-zoster, and HIV. Once the virus has entered the brain, it may cause microglia and astrocyte activation, a massive elevation of cytokines, astrocyte injury, neurodegeneration, and neuronal cell death^18^, contributing to the neuropsychiatric symptoms seen in post-COVID-19 patients^44^. Although we did not observe a relationship between olfactory changes and psychiatric symptoms or cognitive deficits in our preliminary data, with ongoing data collection, we will track longitudinal relationships between changes in olfactory structure, anosmia, and other neuropsychiatric symptoms. Prior work has shown that both OB volume and OS depth abnormalities can persist at 10-12 months after infection^45^.

### Neural Circuit Dysfunction

Our preliminary functional neuroimaging results corrected for multiple testing suggest that post-COVID-19 patients display intrinsic FC dysfunction that has been previously demonstrated in depression and anxiety, including hypoconnectivity in the attention network^46^ and hypoconnectivity between the vmPFC and ventral striatum^47^. Notably, prior work in depressed participants has demonstrated that inflammation is associated with decreased resting-state FC in a widely-distributed network that includes hypoconnectivity between the medial prefrontal cortex and precuneus^13^. Additionally, our finding that functional connectivity between the vmPFC and ventral striatum is reduced in IRIS participants is notable in the context of previous work showing that reduced corticostriatal FC is associated increased inflammation^12, 13^. Our next step will be to explore relationships between levels of inflammatory cytokines and functional neuroimaging to understand potential predictors and mechanisms of neural circuit dysfunction in post-COVID-19 patients.

Our exploratory fMRI results at the voxel-level p < 0.1 (uncorrected) – presented to help guide future research in post-COVID-19 patients – indicate hyperconnectivity in the DMN, which has previously been observed in depression^46^. Furthermore, during the Go-NoGo task in which DLPFC and ACC are typically activated within the cognitive control circuit, the standardized IRIS participants showed hypoactivation in the DLPFC and hyperactivation of the ACC, which has previously been demonstrated in depression during a cognitive task^30^. This pattern may be reflective of overcompensation of the ACC in response to DLPFC underactivity. Hyperactivation in the visual cortex is expected due to the visual nature of the task. We also observed hyperactivation of the left temporal gyrus, which, to our knowledge, has not been observed previously in depressed or anxious participants in response to a cognitive task and should be further explored in the context of long COVID.

The happy vs. neutral contrast of the Viewing of Facial Emotion Task is known to elicit activity in components of the positive affect or reward circuit, including the ventral striatum, orbital frontal cortex, mPFC, and DLPFC. In terms of regions that were differentially activated between IRIS participants and controls at the p < 0.1 level, we observed hypoactivation in the mPFC and hyperactivation in the DLPFC, a pattern which interestingly been observed in recovery from depression^48^. We also found hypoactivation in the PCC and hyperactivation in the bilateral precentral cortex and bilateral middle frontal lobe in response to the happy vs. neutral contrast, findings which have not been shown in response to a rewarding task to our knowledge in depression or anxiety and should be investigated further in relation to COVID-19.

The threatening vs. neutral and sad vs. neutral contrasts are known to evoke activity in the amygdala, insula, and ACC. Notably, we did not observe any differences between IRIS participants and controls in these regions. Instead, IRIS participants showed hyperactivation in the right DLPFC and bilateral middle frontal lobe and hypoactivation in the thalamus and caudate with threatening faces, while they demonstrated hypoactivation in the PCC, mPFC, and left superior frontal lobe with sad faces. Again, these are regions of potential interest in the mechanisms underlying psychiatric symptoms in post-COVID-19 patients.

With our current sample size, we do not observe any discernible relationships between individual intrinsic FC and activation maps and psychiatric symptoms or cognitive performance. There are multiple potential explanations for this, including that our sample size of neuroimaging participants is not large enough yet for patterns to emerge. Another potential explanation is that additional biological, clinical, or environmental factors may need to be considered to explain the relationship between neural function, psychiatric symptoms, and cognitive performance. Finally, the lack of relationship may be due to the fact that the neuroimaging data was collected after the symptom and neurocognitive performance data.

Limitations of the current study include a small sample size for the neuroimaging subsample and the fact that participants’ prior psychiatric diagnoses and current psychotropics were not considered in the analyses. Future analyses with this cohort will include an expanded neuroimaging sample and consideration of these measures. An additional limitation is the variable timeframe in which the first visit occurred after diagnosis due to the challenges of initiating a study during the pandemic. Additionally, the neuroimaging data was collected after the symptom and cognition data. Finally, while we have compared the sample to normative samples collected prior to the pandemic, we do not have data on these individuals prior to developing COVID-19, so we cannot be certain that the deficits we observe were not present prior to contracting SARS-CoV-2.

## Conclusion

With over 200 million patients diagnosed with COVID-19 to date worldwide^49^, post-COVID sequelae are a major public health burden and will continue to be so for many years to come. Prior findings have demonstrated that approximately a third of those infected by SARS-CoV-2 develop mental health problems^40^ with staggering consequences for individuals, families, health care systems, and communities^50^. Furthermore, post-COVID-19 patients demonstrate cognitive deficits on both self-report measures^15^ and objective testing^16^. Our preliminary results support and extend these findings. To our knowledge, our study is the first to examine functional neuroimaging findings in post-COVID participants in conjunction with self-report psychiatric questionnaires and objective neurocognitive testing. We extend the current findings in the literature by demonstrating that psychiatric symptoms, neurocognitive dysfunction, and structural abnormalities are accompanied by dysfunction in several brain networks. In order to develop targeted treatments and prevention strategies for neuropsychiatric sequelae of COVID-19, it will be critical to track profiles of brain structure and function over time in COVID-19 patients and relate them to baseline presentation and risk factors, neuropsychiatric symptoms, neurocognitive performance, and inflammatory and immune profiles.

## Acknowledgements

The authors would like to extend their appreciation to the participants in this study.

## Ethics Approval and Consent to Participate

The Institutional Review Board of Stanford University has approved this protocol (#57070). A study coordinator thoroughly explained the protocol to participants and answered any questions before they provided written informed consent to begin the study. The study was conducted according to the principles of the Declaration of Helsinki.

## Authors’ Contribution

LMH, MW, AS, PG, HB, and LMW developed the study; LMH, JB, XZ, BJ, and LMW performed the data analysis. LMH, JB, and MC wrote the manuscript, and all authors reviewed and approved the manuscript.

## Data Availability Statement

Data collection is ongoing. Data will be made publicly available once data collection has been completed.

