## Supplemental Figures for "Survivors of SARS-CoV-2 Infection Show Neuropsychiatric Sequelae Measured by Surveys, Neurocognitive Testing, and Magnetic Resonance Imaging: Preliminary Results"

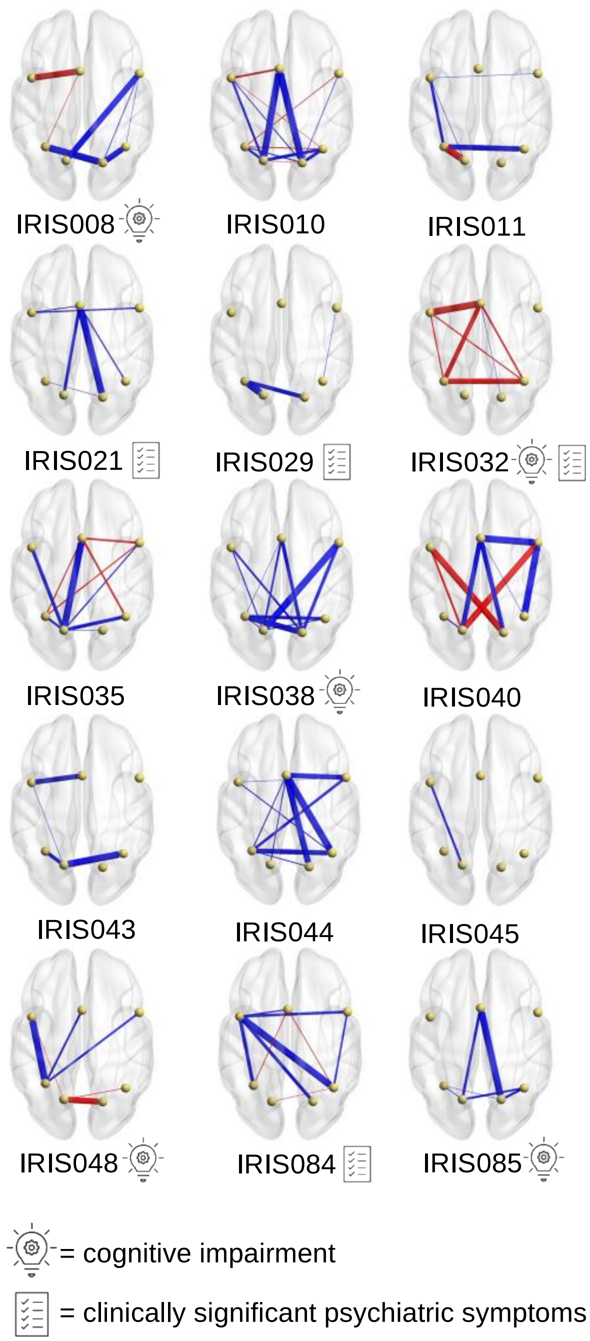


**Supplemental Fig. 1. Individual Intrinsic Functional Connectivity Maps for the Attention Network (z > 1).** Maps are standardized to healthy controls with blue indicating lower intrinsic functional connectivity (FC) and red indicating higher FC as compared to controls. The thickness of the lines represents the strength of FC. Participants with the "cognitive impairment" icon fell within the impaired group in the clustering analysis and those with the "clinically significant psychiatric symptoms" icon had moderate to severe depression and/or anxiety symptoms.

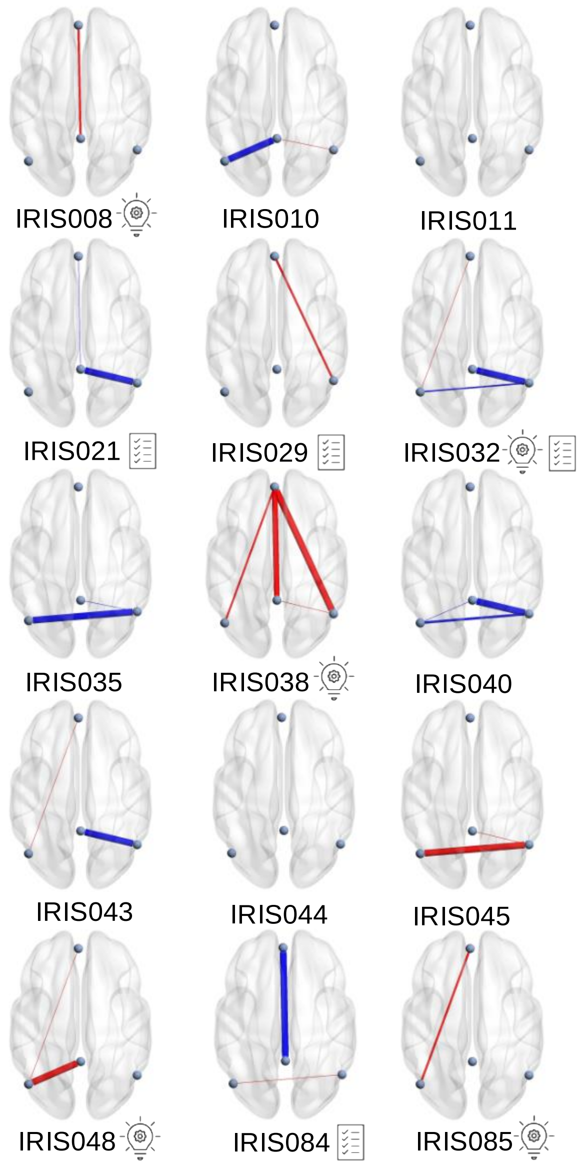


= cognitive impairment

= clinically significant psychiatric symptoms

**Supplemental Fig. 2. Individual Intrinsic Functional Connectivity Maps for the Default Mode Network (z > 1).** Maps are standardized to healthy controls with blue indicating lower intrinsic functional connectivity (FC) and red indicating higher FC as compared to controls. The thickness of the lines represents the strength of FC. Participants with the “cognitive impairment” icon fell within the impaired group in the clustering analysis and those with the “clinically significant psychiatric symptoms” icon had moderate to severe depression and/or anxiety symptoms.

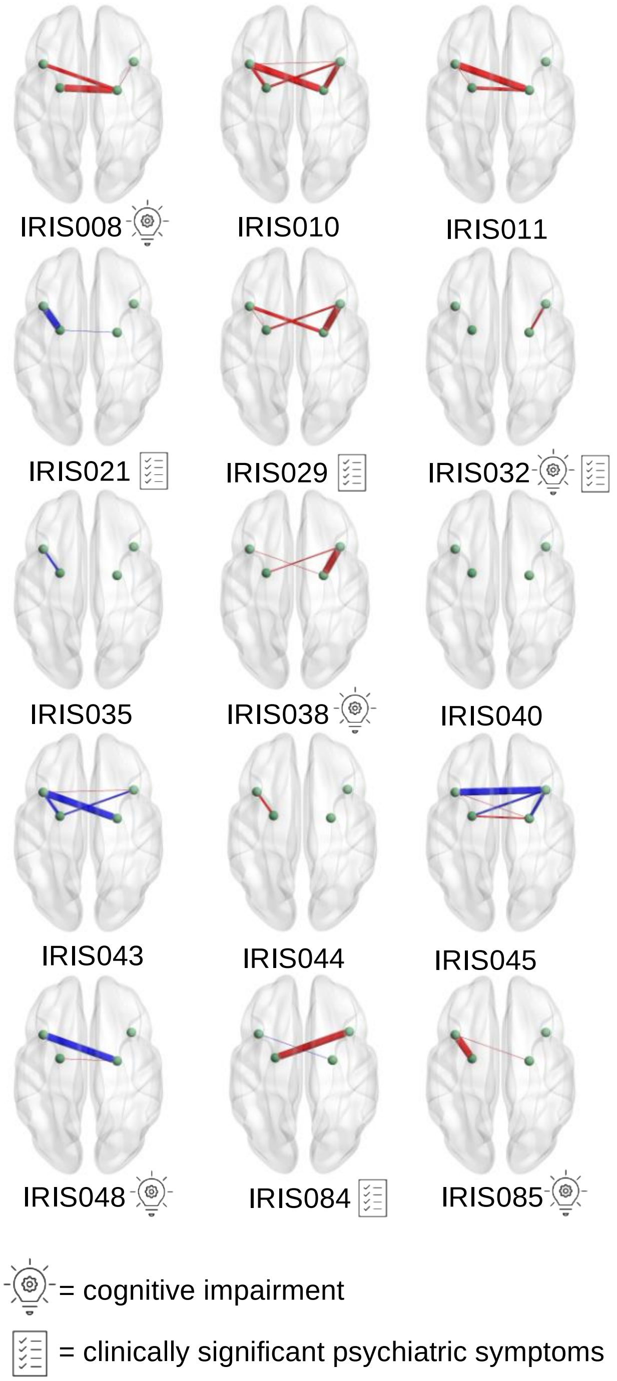


**Supplemental Fig. 3. Individual Intrinsic Functional Connectivity Maps for the Salience Network (z > 1).** Maps are standardized to healthy controls with blue indicating lower intrinsic functional connectivity (FC) and red indicating higher FC as compared to controls. The thickness of the lines represents the strength of FC. Participants with the "cognitive impairment" icon fell within the impaired group in the clustering analysis and those with the "clinically significant psychiatric symptoms" icon had moderate to severe depression and/or anxiety symptoms.

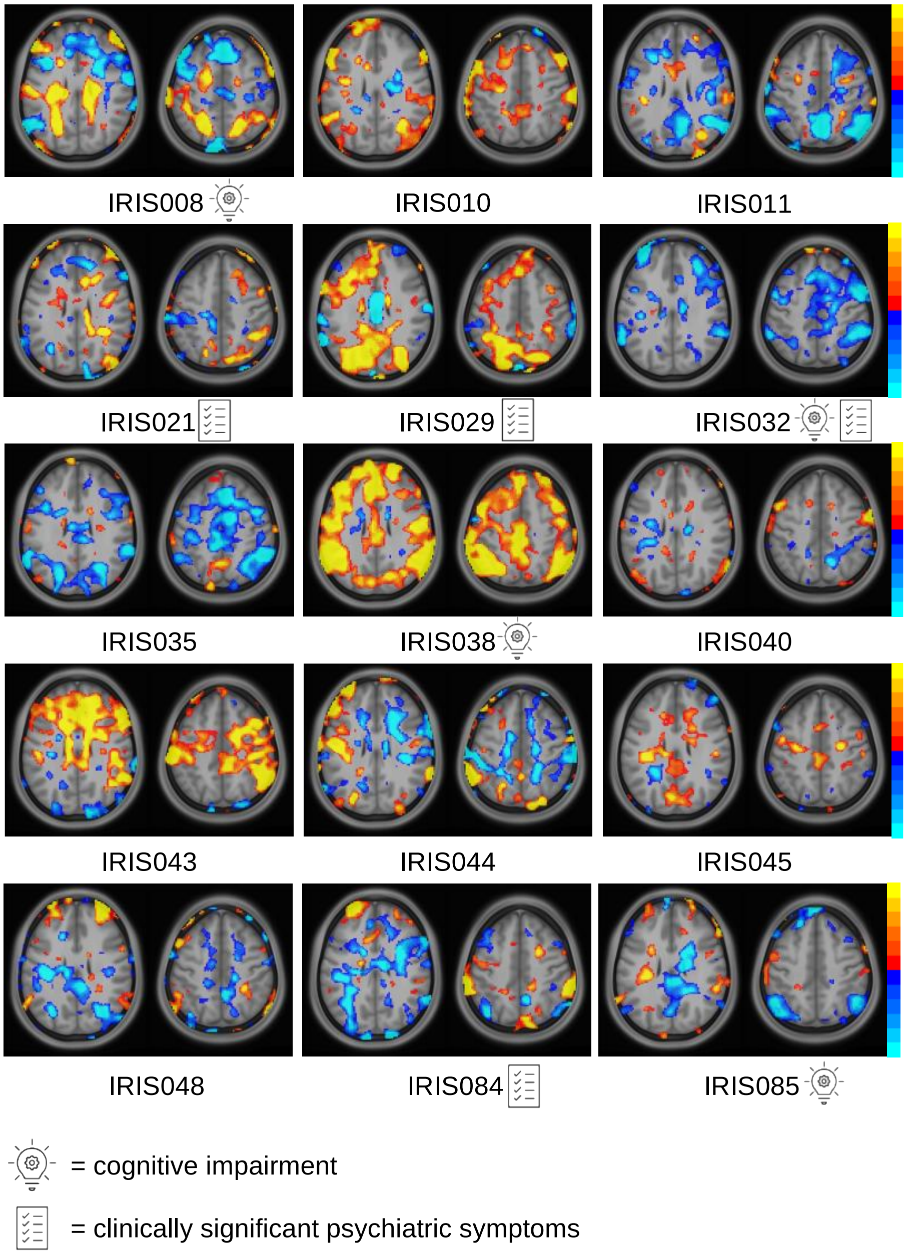


**Supplemental Fig. 4. Individual Intrinsic Functional Connectivity Maps for Seed in the Ventral Medial Prefrontal Cortex (vmPFC) (z > 1).** Maps are standardized to healthy controls with warm colors representing hyperactivation (ranging from 1 to 2) and cool colors representing hypoactivation (ranging from -1 to -2). Participants with the “cognitive impairment” icon fell within the impaired group in the clustering analysis and those with the “clinically significant psychiatric symptoms” icon had moderate to severe depression and/or anxiety symptoms.

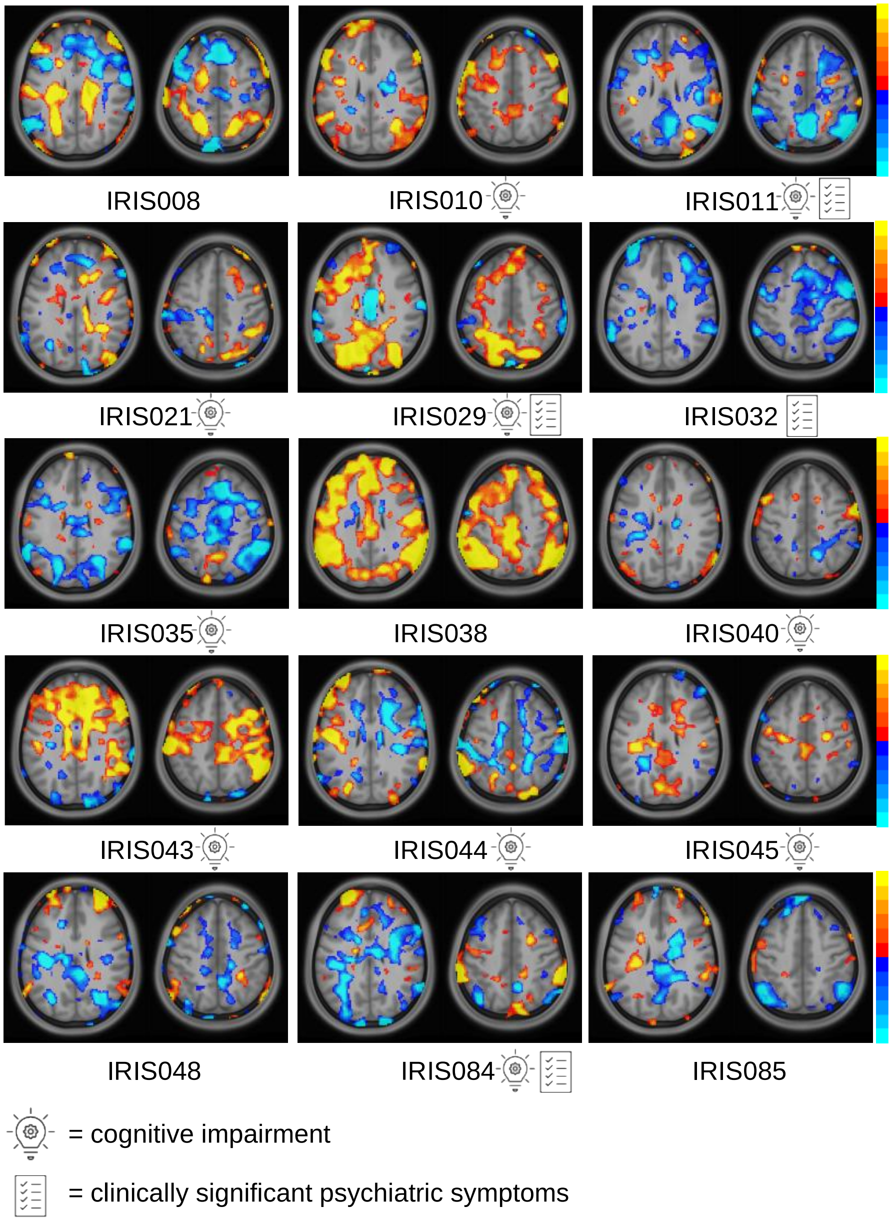


**Supplemental Fig. 5. Individual Intrinsic Functional Connectivity Maps for the GoNoGo Task ($p < 0.3$, $z > 1$).** Maps are standardized to healthy controls with warm colors representing hyperactivation (ranging from 1 to 2) and cool colors representing hypoactivation (ranging from -1 to -2). Participants with the "cognitive impairment" icon fell within the impaired group in the clustering analysis and those with the "clinically significant psychiatric symptoms" icon had moderate to severe depression and/or anxiety symptoms.

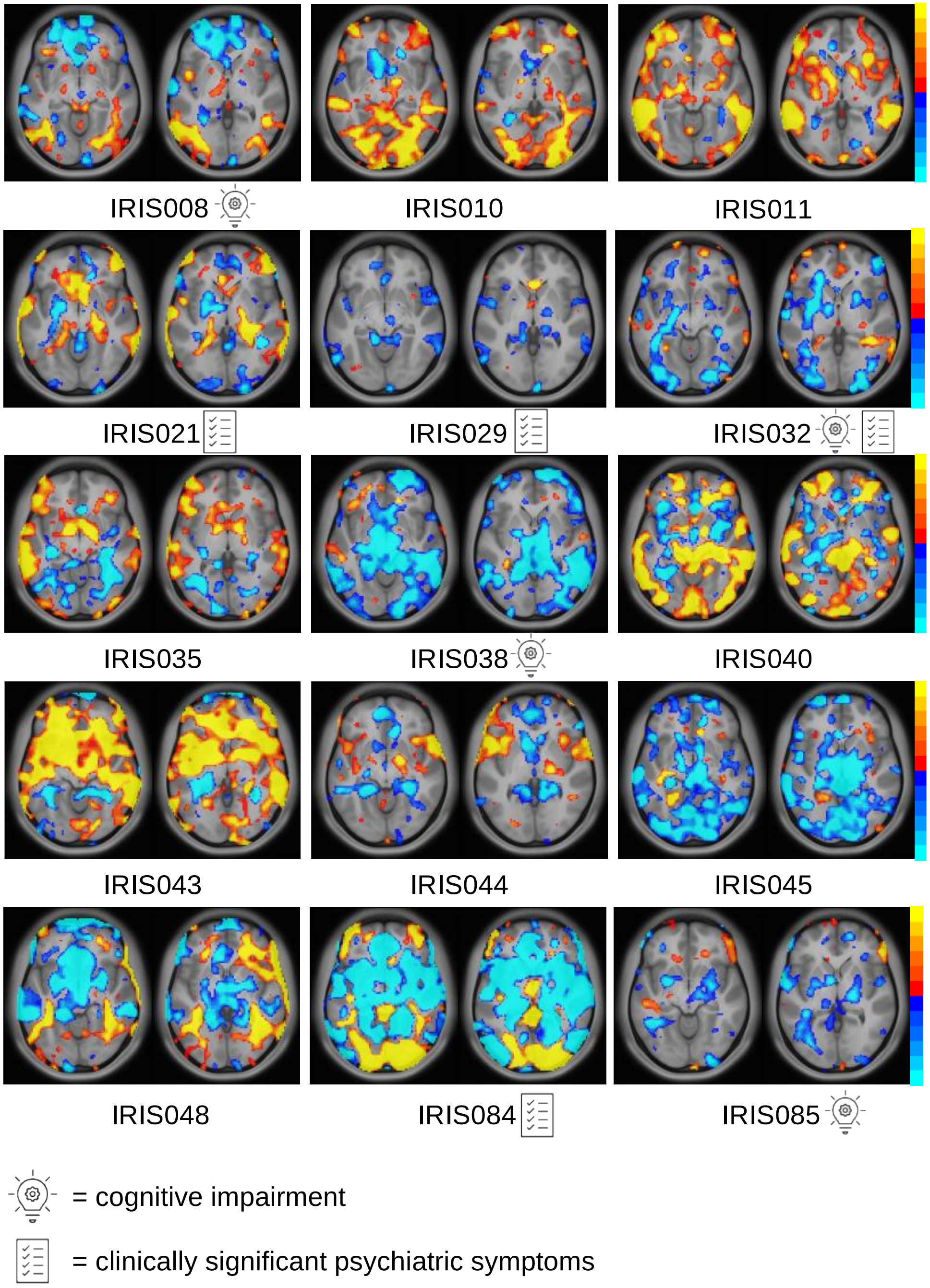


**Supplemental Fig. 6. Individual Activation Maps for Happy Faces (positive affect) Relative to Neutral in the Viewing of Facial Emotion Task (z > 1).** Maps are standardized to healthy controls with warm colors representing hyperactivation (ranging from 1 to 2) and cool colors representing hypoactivation (ranging from -1 to -2). Participants with the "cognitive impairment" icon fell within the impaired group in the clustering analysis and those with the "clinically significant psychiatric symptoms" icon had moderate to severe depression and/or anxiety symptoms.

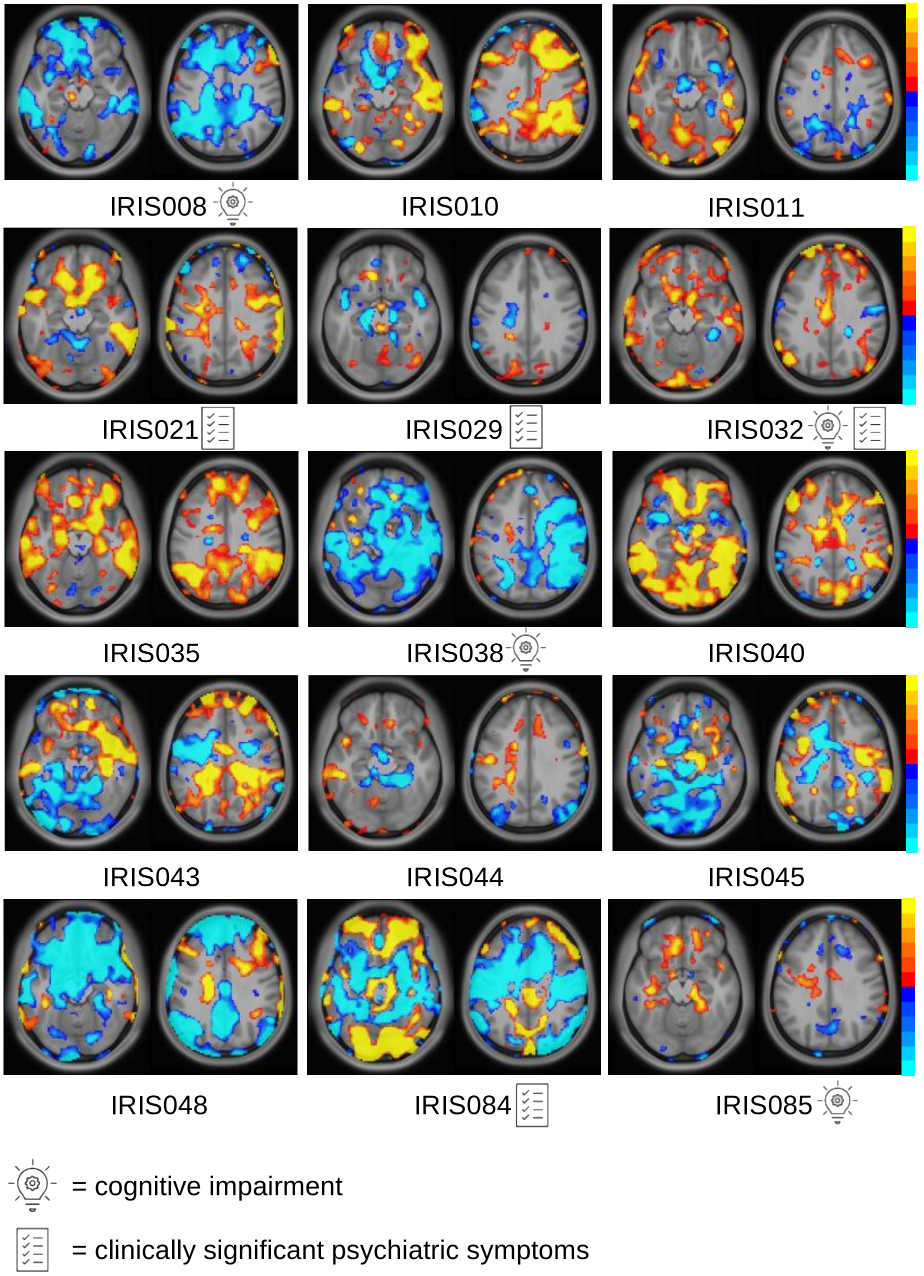


**Supplemental Fig. 7. Individual Activation Maps for Threatening Faces (negative affect) Relative to Neutral in the Viewing of Facial Emotion Task (z > 1).** Maps are standardized to healthy controls with warm colors representing hyperactivation (ranging from 1 to 2) and cool colors representing hypoactivation (ranging from -1 to -2). Participants with the “cognitive impairment” icon fell within the impaired group in the clustering analysis and those with the “clinically significant psychiatric symptoms” icon had moderate to severe depression and/or anxiety symptoms.

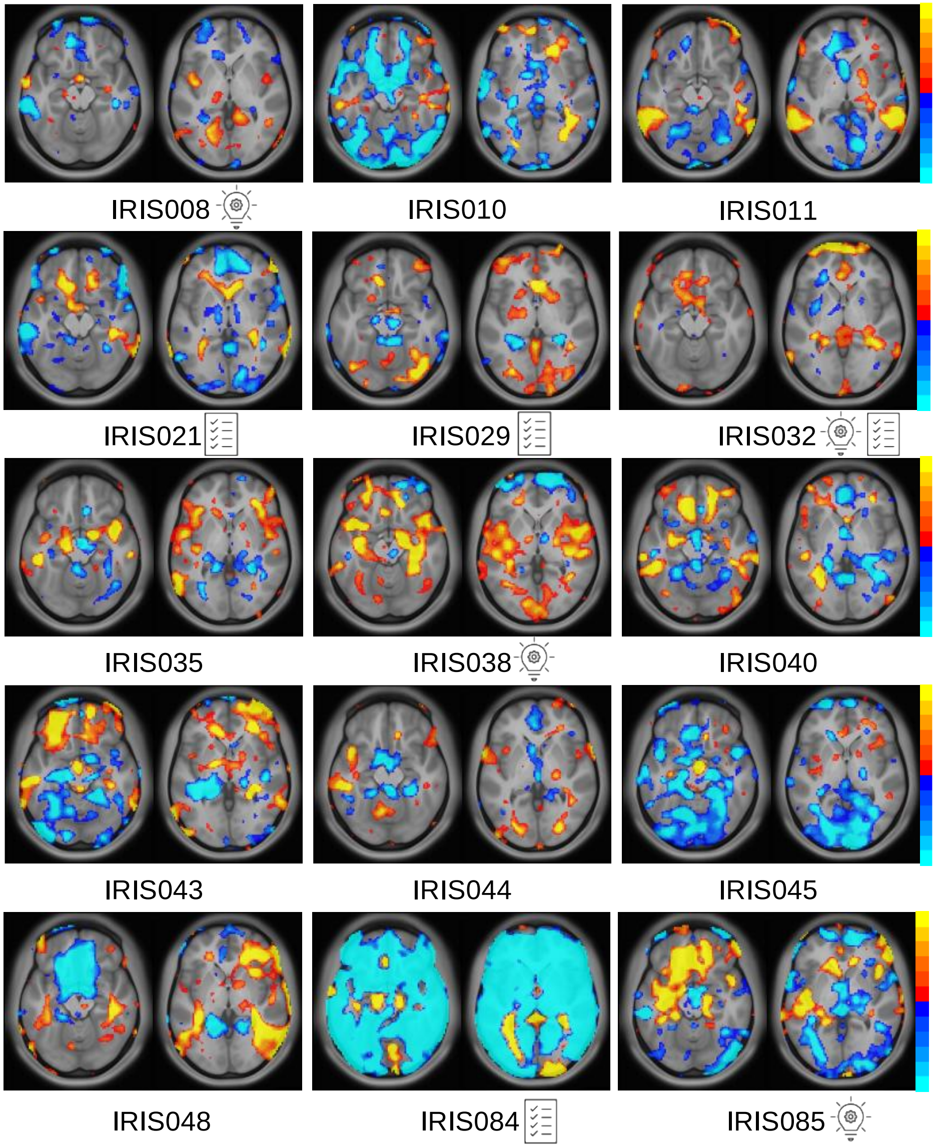


= cognitive impairment

= clinically significant psychiatric symptoms

**Supplemental Fig. 8. Individual Activation Maps for Sad Faces (negative affect) Relative to Neutral in the Viewing of Facial Emotion Task (z > 1).** Maps are standardized to healthy controls with warm colors representing hyperactivation (ranging from 1 to 2) and cool colors representing hypoactivation (ranging from -1 to -2). Participants with the "cognitive impairment" icon fell within the impaired group in the clustering analysis and those with the "clinically significant psychiatric symptoms" icon had moderate to severe depression and/or anxiety symptoms.
